# Neurophysiological resting-state EEG markers of catatonia in schizophrenia and mood disorders

**DOI:** 10.1101/2025.06.26.25327974

**Authors:** Mylène Moyal, Aline Lefebvre, Sahar Allouch, Sophie B. Sebille, David M. Alexander, Laura Dugué, Mahmoud Hassan, Victor Férat, Marie-Odile Krebs, Martine Gavaret, Boris Chaumette, Marion Plaze, Anton Iftimovici

## Abstract

**Key points:** *Question:* Is catatonia, hypothesized to involve disrupted excitation–inhibition balance and cortical dysconnectivity, associated with quantitative abnormalities on resting-state EEG?

*Findings:* In this retrospective, case-control, transdiagnostic hospital-based cohort of patients with schizophrenia and/or mood disorders, we analyzed resting-state EEG data comparing individuals with (n=102) and without (n=519) catatonia. Patients with catatonia showed significant increase in delta power, decrease in alpha power, increase in gamma power, significant reduction in peak alpha frequency and longer mean duration of microstate C.

*Meaning:* Power spectral density, alpha peak frequency, and microstate disruption in catatonia suggest a neurodevelopmentally-related excitation/inhibition dysregulation and support the use of routine clinical EEG for developing diagnostic biomarkers.

**Importance:** Catatonia is a severe psychomotor syndrome that complicates many psychiatric, neurodevelopmental, and non-psychiatric conditions. Identifying reliable diagnostic biomarkers remains a key challenge to improve early intervention and reduce morbimortality. Since its pathophysiology may involve cortical dysconnectivity, electroencephalography (EEG) could provide accessible disease-associated measures, such as power spectral density (PSD - reflecting excitation/inhibition balance), peak alpha frequency (PAF - related to deviations in neurodevelopmental trajectories), and C and D microstates (previously linked respectively to self-referential and externally-oriented attentional modes). However, EEG is yet to be used for this purpose.

**Objective:** To leverage routine clinical EEG recordings to identify neurophysiological markers associated with catatonia. We postulate to find anomalies in PSD distribution, a decrease in peak alpha frequency, and increases in the representation of C over D microstate classes.

**Design:** This study is a case-control retrospective transdiagnostic hospital-based cohort of patients with schizophrenia and/or mood disorders.

**Participants:** We analyzed resting-state EEG data from patients diagnosed with schizophrenia or mood disorders, both with (n=102) and without (n=519) catatonia. EEG data were preprocessed using a well-validated multistep automated pipeline.

**Exposures/measures:** Linear regression models assessed associations between catatonia status and PSD, PAF and microstates, adjusting for age, sex, medication (computed as olanzapine, fluoxetine, and diazepam equivalents), and comorbid neurodevelopmental or neurological conditions.

**Results:** Patients with catatonia showed increased delta power (T = 2.37, *p*_FDR_ = 0.03), decreased alpha power (T = –3.55, *p*_FDR_ = 0.002) and increased gamma power (T = 3.14, *p*_FDR_ = 0.008), reduced PAF (T = –2.60, *p* = 0.03), and longer mean duration of microstate C (T = 2.17, *p* = 0.03). Results were consistent in the subgroup not receiving benzodiazepines.

**Conclusion and relevance:** Routine clinical EEG revealed quantitative neurophysiological differences between patients with and without catatonia in a transdiagnostic population with psychotic and mood disorders. Power spectral density, alpha peak frequency and microstate anomalies in catatonia shed light on its underlying pathophysiology, suggesting a probable neurodevelopmentally-related excitation/inhibition dysregulation. Importantly, this indicates that routine clinical EEG could be used for diagnostic biomarker development, which would ultimately improve early detection and treatment.

## INTRODUCTION

Catatonia is a severe psychomotor syndrome associated with mood, psychotic, and neurodevelopmental disorders, as well as non-psychiatric medical conditions^1^. Its clinical heterogeneity leads to underdiagnosis^2^ and significant morbimortality^1,3^ despite established DSM5 criteria and a well-identified symptomatic treatment that includes lorazepam and/or electroconvulsive therapy^3,4^. Identifying diagnostic biomarkers is therefore a critical challenge that must be addressed to improve early intervention and reduce the burden of disease.

The pathophysiology of catatonia remains poorly understood. At the molecular level, catatonia is hypothesized to stem from an imbalance between cortical excitatory and inhibitory activity, involving glutamatergic and GABAergic neurotransmission^5,6^. This hypothesis is supported by the efficacy of lorazepam^7,8^ and clozapine^9^. Lorazepam enhances GABA binding at GABA_A_ receptors, while clozapine may facilitate binding at GABA_B_ receptors^10^. These pharmacological effects point to impaired tonic GABA transmission and a complex GABA_A_–GABA_B_ interaction in catatonia^11,12^. Single-photon emission computed tomography (SPECT) also shows reduced GABA_A_ receptor density in the left sensorimotor cortex in catatonia ^13^, along with GABAergic deficits in the orbitofrontal cortex during negative emotional processing^14^. Additionally, genetic syndromes linked to catatonia involve genes expressed in GABAergic neurons^15^. From an immunological perspective, the frequent occurrence of catatonia in N-methyl-D-aspartate receptor (NMDAR) encephalitis also suggests a role for glutamatergic dysfunction^16^. Animal models further implicate dysregulation of dopaminergic, GABAergic, and glutamatergic neurotransmission in the development of catatonia^17^, while brain magnetic resonance imaging (MRI) reveals abnormal functional connectivity in patients with catatonia within cortico-striato-thalamic, cortico-cerebellar, anterior cingulate-medial orbitofrontal, and lateral orbitofrontal networks^18–24^.

Collectively, this evidence indicates disruption in glutamatergic and GABAergic neurotransmission in catatonia, which may manifest as detectable abnormalities in cortical activity. As such, electroencephalography (EEG) could provide an accessible, non-invasive means to quantify anomalies in cortical activity that could be relevant for catatonia diagnosis. Patients with catatonia routinely undergo EEG in clinical practice as part of the initial differential diagnosis to rule out epilepsy^25^ but it can also provide quantitative markers of the temporal dynamics of neuronal networks^26^, and neuronal excitation/inhibition imbalance^27^. To date, however, no published studies have investigated quantitative EEG measures in catatonia.

Here, we studied three types of quantitative EEG markers in relation to catatonia: power spectral density in each frequency band, alpha peak frequency, and microstates. 1) *Power Spectral Density* (PSD) reflects the distribution of power across frequency bands. Slow-frequency rhythms—delta (0.5–4 Hz), theta (4–8 Hz), and alpha (8–12 Hz)—are associated with long-range connectivity and play a role in synaptic plasticity and memory processes^28^. In contrast, fast-frequency rhythms—beta (12–30 Hz) and gamma (30–45 Hz)— support short-range connectivity and are linked to higher-order cognitive functions such as attention and conscious perception^28^. In addition, beta and gamma rhythms were shown to rely on an interplay between excitatory and inhibitory neuronal activity^27^. Disruption in all frequency bands has been reported across psychiatric disorders^29^. 2) *Peak alpha frequency* (PAF), defined as the frequency of the maximum power within the alpha range, is reduced across neurodevelopmental and schizophrenia-spectrum disorders^30,31^. 3) *Microstates* are transient, global patterns of polarization of the electric field on the scalp that fluctuate on a millisecond timescale^32^. They have been associated with GABA-R availability^33^ and are thought to represent the electrophysiological counterparts of resting-state networks identified by functional magnetic resonance imaging (fMRI), but at a higher temporal resolution^34–38^. Specifically, an increase in class C microstates (associated with self-referential and interoceptive processing) and a decrease in class D microstates (linked to externally directed attentional processes) have been observed across neurodevelopmental and schizophrenia-spectrum disorders^39–43^.

We postulate that a severe condition such as catatonia results from a disruption in excitation/inhibition balance more important than the one already expected in schizophrenia and mood disorders, reflected by anomalies in PSD distribution, a decrease in peak alpha frequency, and an increase in the representation of C over D microstate classes. To test the relevance of these markers in a real-world setting, we compared them between patients with and without catatonia (respectively N=102 and N=519), extracted from a transdiagnostic retrospective hospital-based cohort of patients with schizophrenia and/or mood disorders.

## 1. METHODS

### 1.1 Participants

We screened all individuals who received a diagnosis of schizophrenia or mood spectrum disorders (based on ICD-10 diagnostic codes) and who also underwent EEG recordings between 2019 and 2024, extracted from the GHU Paris Psychiatry and Neurosciences data warehouse (N = 850). To identify patients with catatonia, we used an automated textual research looking for the expression “catato*” or searched for ICD-10 diagnostic codes F061 “organic catatonia” or F20.2 “schizophrenic catatonia”. Each of the resulting medical records was then manually checked to confirm the diagnosis of catatonia according to DSM-5 criteria (i.e., three criteria among catalepsy, stupor, waxy flexibility, agitation, mutism, negativism, posturing, mannerisms, stereotypies, grimacing, and echo phenomena). The remaining patients were included in the non-catatonia group. The following information was extracted for every patient: age at the time of the EEG, sex, underlying diagnosis, presence of neurological or neurodevelopmental comorbidities (see Supplementary data S1 for the list of ICD codes), and ongoing treatment when the EEG was undertaken, converted into equivalents of olanzapine, diazepam and fluoxetine. Patients without all the available phenotypic data were excluded (N = 120). This study was approved by the GHU Paris Research Ethics Committee on February / 06 / 2024 (agreement number 2024-CER-A-010 – TEMPO project).

### 1.2 EEG recording

EEG data were recorded using 21 scalp electrodes with gel, following the international 10–20 system^44^, on a Micromed® set-up. Impedances were kept below 5 kΩ. Sampling rate was 256 Hz. Routine clinical EEGs had a mean duration of 20 min. They were performed at rest in a half-seated position, and included rest sequences with eyes closed and eyes open, followed by stimulation procedures (hyperpnea and then intermittent photic stimulation), all of which were annotated. Patients without annotated EEG were excluded (N=31).

### 1.3 EEG preprocessing

Preprocessing was done using the MNE-Python software^45,46^, according to the OHBM COBIDAS MEEG good practice recommendations^47^ (full code is available on github.com/mylenemoyal/Catatonia-EEG-PSD-microstates). The EEG preprocessing utilized a multi-step automated algorithm employed in recent studies^48,49^. EEG signals underwent bandpass filtering (1-100 Hz). Bad EEG channels were identified using the pyprep algorithm, which employs a RANSAC-based method, and then interpolated using spherical spline interpolation from neighboring good channels^50^. Electrodes were then re-referenced to the common average. Artefacts were detected using independent component analysis (ICA) combined with the IClabel algorithm^51,52^. ICA decomposition was performed with the number of components set to the number of EEG channels (i.e., 19). To account for the high level of noise associated with this clinical data, we set stringent thresholds to keep components labeled as “brain-related” (> 70%) and reject those labeled as artefacts (> 80%). A second bandpass filter (1-45 Hz) was then applied. Only initial sequences at rest with eyes closed and before stimulation procedures were used in the main analysis (∼7 min). Next, EEG signals were segmented into 2-second epochs. The Autoreject toolbox^53^ was used to interpolate or reject bad epochs. If there were three or more interpolated channels or two interpolated channels that encompassed the entire occipital region, the participant was excluded from the analysis (N=78). A total of 621 individuals passed these quality checks including 102 with and 519 without catatonia (**Figure 1**). Their demographic and clinical characteristics are reported in **Table 1**. For each EEG recording, the following preprocessing criteria were extracted: channel interpolation number (number of interpolated channels out of the total number of channels), ICA rejection rate (number of ICA components rejected out of the 19 generated) and epoch rejection rate (number of epochs rejected out of the total number of epochs) (**Table 1**).

**Figure 1.**
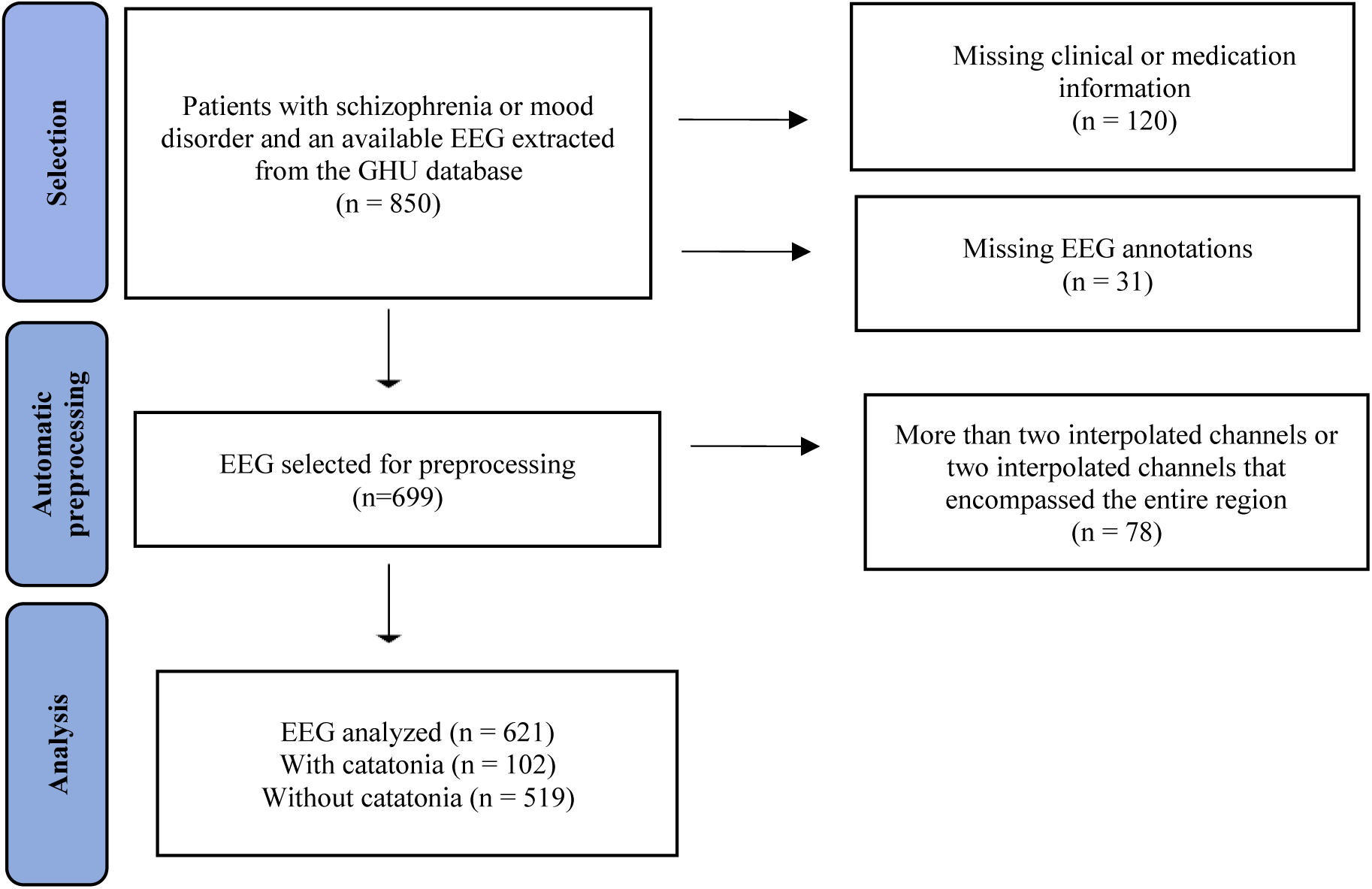
Flow chart.

**Table 1.** Description of the study population. Bold font indicates statistically significant effects. Comparisons were made using either the Z-score proportion test or the Chi^2^, as appropriate. Results are presented as mean ± standard deviation (M ± SD) or as absolute numbers (N) with corresponding percentages.

|  |  | Diagnostic groups |  | Tests |  |
| --- | --- | --- | --- | --- | --- |
| Demographic | N=621 | Catatonia (N=102) | Without catatonia (N=519) | X<sup>2</sup> or Z-score | p |
|  | Age at EEG, M ± SD | 47.5 ± 19 | 41.7 ± 18.2 | 2.83 | **0.005** |
|  | Sex (F/M) | 58(57) / 44(43) | 229(44) / 290(56) | -2.35 | **0.01** |
| Diagnoses | Psychotic disorder N(%) | 59(58) | 357(69) | 2.19 | **0.02** |
|  | Mood disorder N(%)<br>Including Bipolar disorder N(%) | 54(53)<br>22(21) | 253(48)<br>174(33) | -0.7<br>2.38 | 0.4<br>**0.01** |
|  | Neurodevelopmental disorder N(%) | 17(16) | 39(7) | -2.95 | **0.003** |
|  | Neurologic disorder N(%) | 15(15) | 31(6) | -2.89 | **0.003** |
| Treatment | Olanzapine equivalent dosage (mg), M± SD | 27.7 ± 39 | 32.1 ± 52 | -0.975 | 0.33 |
|  | Diazepam equivalent dosage (10mg), M± SD | 10.8 ± 32 | 5.9 ± 30 | 1.419 | **0.05** |
|  | Fluoxetine equivalent dosage (40 mg), M± SD | 0.37 ± 0.66 | 0.29 ± 0.65 | 1.102 | 0.271 |
| Preprocessing criteria | Channel interpolated rate<br>M ± SD | 0.9 ± 0.6 | 0.9 ± 0.7 | 0.132 | 0.895 |
|  | ICA rejection rate<br>M ± SD (%) | 77± 15 | 73 ± 14 | 2.44 | **0.015** |
|  | Epoch rejection rate<br>M ± SD (%) | 2.6 ± 5 | 3.5 ± 7 | 1.41 | 0.15 |

### 1.4 EEG Quantitative Features Analysis

Overall PSD was extracted using Welch’s method on 2-second epochs^54^. For each frequency band, relative power was obtained by averaging power across the channels and within the respective band and normalizing by the total broadband power (**Figure 2**). Peak alpha frequency (PAF) was computed as the frequency of the maximum power within the alpha range (8-12Hz) (**Figure 3**). Both relative power and PAF were averaged across channels to yield one value per subject.

**Figure 2.**
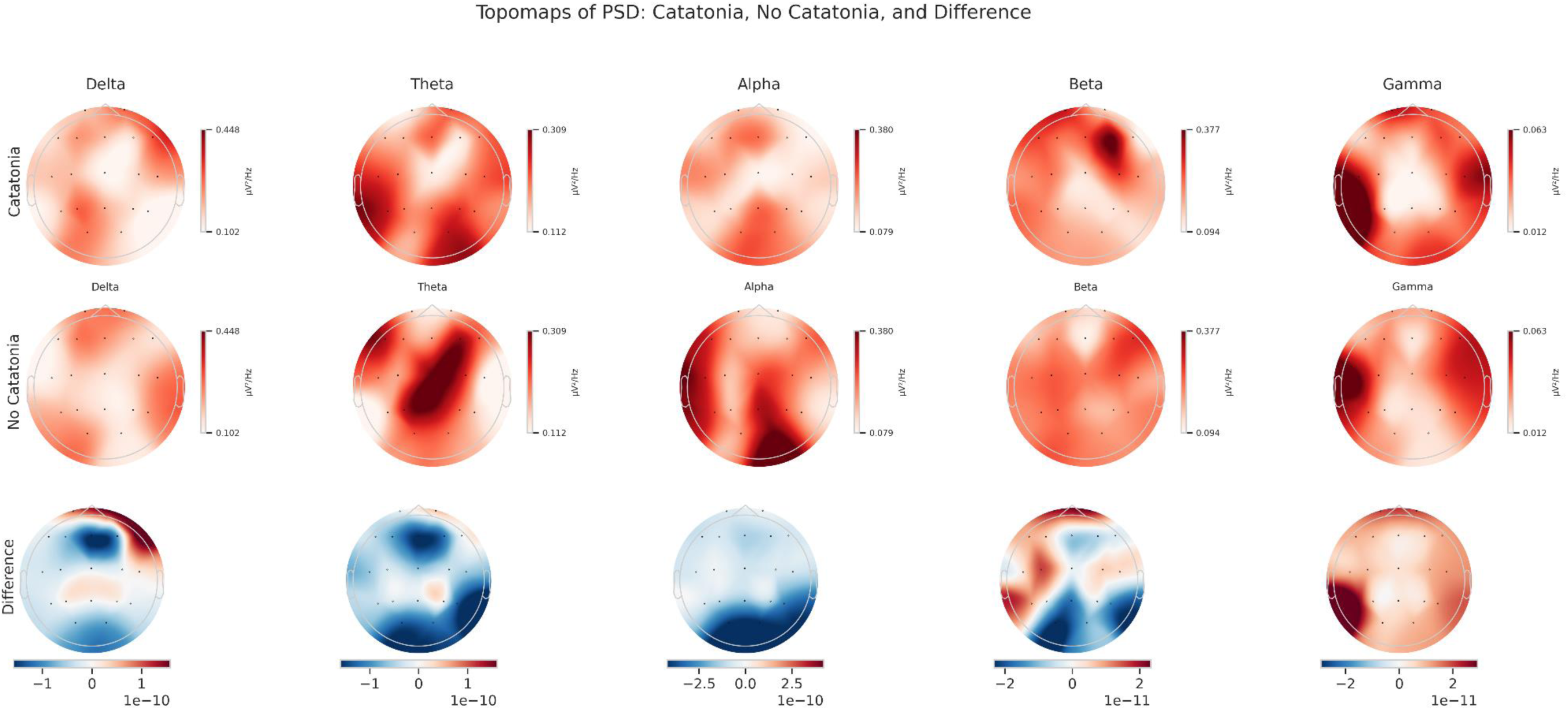
Topographic maps illustrating the spatial distribution of power spectral density (PSD) across frequency bands. The top row displays data from catatonia patients, while the second row shows data from non-catatonia patients. The bottom row presents the relative power difference between the two groups, with blue (red) indicating lower (higher) power values in the catatonia group.

**Figure 3.**
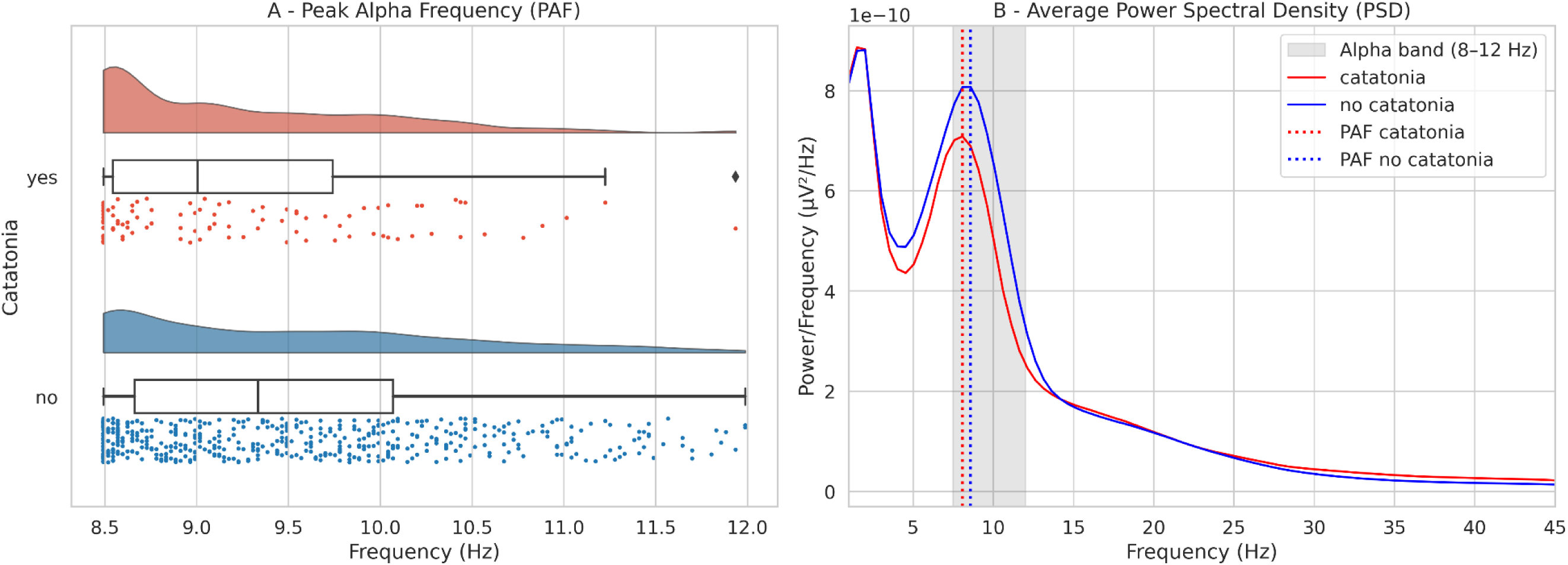
**A**. Peak alpha frequency by group with catatonia patients in red and non-catatonia patient in blue. The rain cloud plots show the estimated probability density of the alpha frequency where the power is maximal, a box plot (the edges of the box are the interquartile range, the central mark of the box is the median, and the whiskers are 1.5 times the interquartile range), and a histogram with data points by individual. **B.** Power spectrum from 0 to 45 Hz. The shaded area reflects a range of 8 to 12 Hz. Mean PSD as a function of frequency (Hz) for catatonia patients (red) and non-catatonia patients (blue). Dotted lines indicate the peak alpha frequency for each group (red for catatonia, blue for non-catatonia).

Microstate analysis was conducted using the Pycrostates library (<u>github.com/vferat/pycrostates</u>)^55^ in the alpha band (8-12Hz) given the resting-state eyes-closed design of our study^56^. Microstate template maps were built in a randomly selected subset of fifty patients without catatonia (see **Supplementary Material S2**). Global field power (GFP) was extracted, and only EEG topographies at GFP peaks were retained for microstate identification using a modified K-means clustering approach. The optimal number of clusters was set to six based on a consensus between the silhouette, Calinski-Harabasz, Davies-Bouldin, and Dunn scores (see **Supplementary material S2**). The topographies were assigned to microstate classes A, B, C, D, E, and F, and backfitted to the rest of the dataset (N=571) (**Figure 4**). For each individual, three parameters were computed for each microstate class: mean duration (average uninterrupted time a given microstate was present, in milliseconds), time coverage (proportion of the total time spent in a given microstate), and occurrence (average number of times a given microstate appeared per second, in Hz).

**Figure 4.**
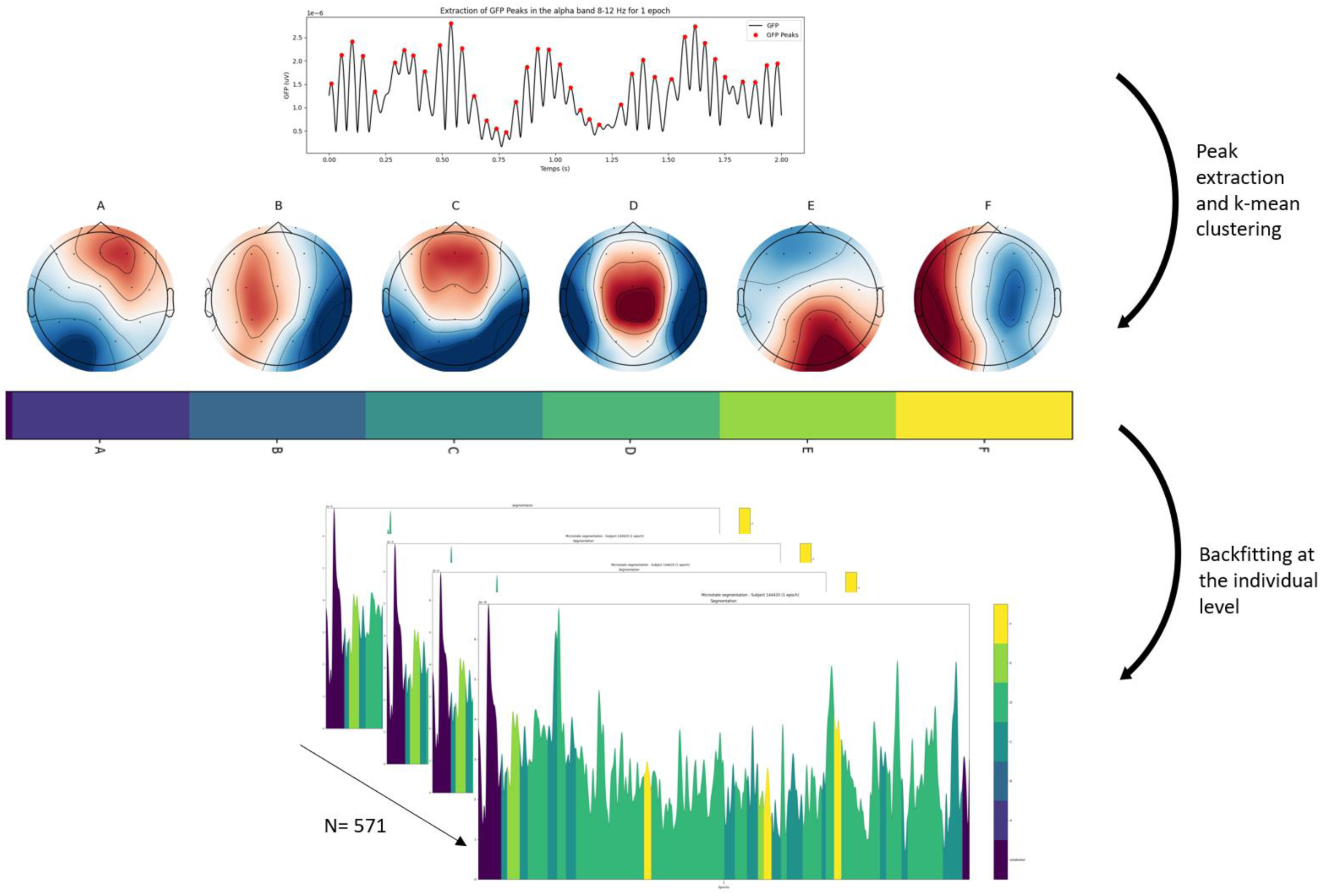
Microstates maps derived from a random subset of 50 patients without catatonia and backfitted to the rest of the patients with and without catatonia.

### 1.5 Statistical analysis

For each feature extracted from the resting-state EEG, we performed a linear regression analysis on catatonia status with the following covariables: age, sex, treatment (total equivalent doses of diazepam, olanzapine, and fluoxetine, which were not correlated to each other – See **Supplementary material S3**), and associated diagnoses (i.e., neurodevelopmental and neurological disorders). In the PSD analysis, results were corrected for multiple comparisons across the five frequency bands using the false discovery rate (FDR) method. PAF and microstate C and D tests were reported with a significance threshold of 0.05, because they were based on a strong preliminary hypothesis. A subgroup analysis was conducted in patients stratified based on their specific psychiatric diagnoses: psychotic disorders (N=417; 59 patients with catatonia versus 358 without catatonia) and mood disorders (N=184; 33 patients with catatonia versus 151 without catatonia). To further control for the effects of benzodiazepines on EEG, we repeated the analysis in a subgroup of patients without benzodiazepines (N = 349; 51 patients with catatonia versus 298 without catatonia – see **Supplementary material S4**).

## 3. RESULTS

### 3.1 Demographic and clinical characteristics

A total of 621 individuals with EEG recordings were included and analyzed. Of these, 102 had catatonia during the EEG recording, while 519 had never been diagnosed with catatonia (**Figure 1** and **Table 1**). Patients with catatonia were older than those without catatonia (47.5 ± 19 vs. 41.7 ± 18.2 years; Z-score = 2.83, p = 0.005), more likely to be women (57% vs. 44%; X^2^= −2.35, p = 0.01), had a lower prevalence of psychotic disorders (58% vs. 69%; X^2^ = 2.19, p = 0.02), and a higher prevalence of neurodevelopmental disorders (16% vs. 7%; X^2^ = −2.95, p = 0.003) and neurological disorders (15% vs. 6%; X^2^ = −2.89, p = 0.003). No significant differences were found for mood disorders. Regarding treatment at the time of EEG, patients with catatonia received higher doses of diazepam equivalents (10.8 vs. 5.9 Z-score=1.419, p=0.05). No significant differences were observed in olanzapine or fluoxetine equivalent dosages.

### 3.2 Power Spectral Density

The overall distribution of topomaps (**Figure 2**) were aligned with physiological expectations, showing predominant occipital activity in the alpha band, and midline-centered theta activity^54,57^

Patients with catatonia exhibited increased power in the delta band (T = 2.37, p_FDR_ = 0.03), decreased power in the alpha band (T = −3.55, p_FDR_ = 0.002), and increased power in the gamma band (T = 3.14, p_FDR_ = 0.008) compared to non-catatonia patients (**Figure 2** and **Table 2**), after correction for age, sex, medication (total equivalent doses of diazepam, olanzapine, and fluoxetine), and associated neurological and neurodevelopmental diagnoses. No significant differences were observed in other frequency bands (see **Supplementary Materials S5**).

**Table 2.** Results of linear regression model between EEG features and catatonia status with covariates of age, sex, treatment (total equivalent of diazepam, olanzapine and fluoxetine), associated diagnosis (neurodevelopmental or neurological disorder). Abbreviations: Bold font corresponds to significant effects. Coef: coefficient; SD: standard deviation; CI[2.5%] CI[97.5%] to the confidence intervals at 2.5 and 97.5% and pval for p.value.

| EEG features | Mean ± SD in catatonia | Mean ± SD in non catatonia | Coef ± SD | T | CI[2.5%]; CI[97.5%] | pval |
| --- | --- | --- | --- | --- | --- | --- |
| **Delta** | **0.039±0.019** | **0.037±0.017** | **0.005±0.002** | **2.374** | **0.001; 0.008** | **0.03\*** |
| Theta | 0.024±0.012 | 0.026±0.013 | 0±0.001 | 0.019 | -0.003; 0.003 | 0.999* |
| **Alpha** | **0.026±0.011** | **0.031±0.013** | **-0.005±0.001** | **-3.553** | **-0.008; -0.002** | **0.002\*** |
| Beta | 0.008±0.003 | 0.008±0.003 | 0±0 | 0.021 | -0.001; 0.001 | 0.999* |
| **Gamma** | **0.002±0.002** | **0.002±0.001** | **0±0** | **3.146** | **0; 0.001** | **0.008\*** |
| **Alpha peak** | **9.207±0.692** | **9.437±0.883** | **-0.248±0.095** | **-2.603** | **-0.435; -0.061** | **0.009** |
| C occurrence | 2.974±1.82 | 2.915±1.802 | 0.046±0.204 | 0.226 | -0.354; 0.446 | 0.821 |
| C time coverage | 0.148±0.127 | 0.135±0.108 | 0.012±0.013 | 0.967 | -0.013; 0.037 | 0.334 |
| **C mean duration** | **0.044±0.014** | **0.042±0.009** | **0.003±0.001** | **2.176** | **0; 0.005** | **0.03** |
| D occurrence | 3.808±1.705 | 3.943±1.777 | -0.076±0.199 | -0.383 | -0.466; 0.314 | 0.702 |
| D time coverage | 0.239±0.191 | 0.258±0.213 | -0.016±0.024 | -0.677 | -0.063; 0.03 | 0.498 |
| D mean duration | 0.059±0.047 | 0.06±0.045 | -0.001±0.005 | -0.274 | -0.011; 0.009 | 0.784 |

In the subgroup of patients without benzodiazepines (N=349), the increase in gamma power remained significant (T = 3.08, p_FDR_ = 0.011 - see **Supplementary Material S8**). The relative distribution of other frequencies was also preserved despite a lack of significance after correction, with a decrease in alpha power (T = −2.11, p_FDR_ = 0.088) and an increase in delta power (T = 1.09, p_FDR_ = 0.458).

In the subgroup analysis by diagnosis, catatonia patients with psychosis exhibited reduced alpha power compared to non-catatonia patients with psychosis (T = −2.63, p_FDR_ = 0.044) (see **Supplementary Material S6**). No significant differences were observed in the mood disorder subgroup (see **Supplementary Material S7**).

### 3.1 Peak Alpha Frequency

Using the same linear regression model, catatonia patients exhibited a reduced peak alpha frequency (T=-2.60, p= 0.009) compared to non-catatonia patients see (**Figure 3** and **Table 2**). This was also the case in the subgroup of patients without benzodiazepines (T = −2.46, p = 0.014). In the subgroup analysis by diagnosis, catatonia patients with psychosis showed a reduced PAF (T = −2.58, p = 0.01), which was not the case for catatonia patients in the mood disorder subgroup (see **Supplementary Material S6 and S7**).

### 3.2 Microstates

The global explained variance (GEV) of the microstates maps extracted from the 50 randomly selected patients was 60%. Topographies were consistent with the literature^34^ (**Figure 4)**. Catatonia patients exhibited a higher mean duration of microstate C than non-catatonia patients (T=2.17, p =0.03), after correction for age, sex, medication (total equivalent doses of diazepam, olanzapine, and fluoxetine), and associated neurological and neurodevelopmental diagnoses (**Table 2**). In the subgroup of patients without benzodiazepines, this increase in class C mean duration remained significant (T = 2.99, p = 0.003), and we also observed a significant increase in class C time coverage in catatonia (T = 1.97, p = 0.049). No differences concerning the microstate C or D were found in the subgroup with psychosis or mood disorder (see **Supplementary Material S6 and S7**).

## DISCUSSION

In this case-control study, quantitative measures of resting-state EEG were compared for the first time between patients with and without catatonia from a large transdiagnostic hospital-based population with schizophrenia and mood disorders. Using routine clinical EEG, we identified significant differences in power spectral densities, peak alpha frequency, and microstate class C between the groups.

Firstly, we observed an increase in delta power and a decrease in alpha power in catatonia at rest. The increase in delta power aligns with previous qualitative EEG studies in this population, which reported a generalized slowing of brain activity^58,59^. An elevated delta/alpha ratio has been documented in psychotic disorders^60,61^ and may reflect the involvement of thalamo-cortical mechanisms^61^. Specifically, thalamic nuclei are known to modulate delta synchronization via GABAergic projections^62^. Impairments in thalamo-cortical connectivity could therefore underlie the observed increase in delta activity^63^. This aligns with the hypothesis of a disruption in cingulate-basal ganglio-thalamo-cortical loop in catatonia^64^ where both abnormal functional thalamocortical connectivity^20^ and structural alterations in the thalamus have been reported^65^.

In addition, the relative distribution of PSD across frequency bands at rest evolves with age, shifting from dominant low-frequency bands in childhood to alpha dominance in adulthood. This shift reflects, from a functional perspective, an increased inhibitory tone in the excitation/inhibition balance^66^, which structurally for alpha band, may be related to the maturation of white matter tracts^67^. The observed reduction in alpha power in individuals with catatonia could therefore reflect a disruption in the development of this inhibitory tone. Moreover, the reduction in peak alpha frequency (PAF) observed in this cohort reinforces this hypothesis of a disrupted neurodevelopmental trajectory: higher PAF is associated with better cognitive abilities^30,31^, it physiologically increases during development, and lower PAF values have previously been associated with neurodevelopmental disorders^31,68,69^. This link between atypical neurodevelopment and the onset of catatonia is further supported by its high prevalence in individuals with neurodevelopmental disorders^15,70^, and by brain imaging studies indicating atypical brain development—deviations in sulcation and gyrification indices in patients with catatonia^65,71^. In this cohort, also, neurodevelopmental disorders were more represented in the catatonia group. Interestingly, both alpha power and PAF were reduced in the psychosis but not the mood disorder subgroups, possibly further reflecting the neurodevelopmental burden that is more present in schizophrenia than in affective disorders^72^.

Secondly, we observed an increase in the lower bands of resting-state gamma power in patients with catatonia. The elevation of resting-state gamma activity is consistent with the hypothesis of an imbalance between cortical excitatory and inhibitory activity in catatonia. Indeed, the enhancement of gamma power has been linked to GABAergic activity, as measured by proton magnetic resonance spectroscopy or pharmacologically induced by propofol, a GABAergic agent^27^. Moreover, NMDA receptor antagonists lead to an increase in power and connectivity in the gamma band in human and preclinical studies^73^. An increase in resting-state gamma activity may also reflect a reduced capacity to further elevate gamma activity in response to stimuli. This is consistent with previous studies suggesting a decrease in the signal-to-noise ratio in schizophrenia, which may impair the generation of task-related oscillations^74,75^. Furthermore, a reduction in cortical excitatory responses—assessed via the short-interval intracortical inhibition (SICI) paradigm, which is closely tied to GABAergic interneuron function^76^—has been associated with general psychomotor slowing in psychosis, including in individuals with catatonia^77^. The balance between excitation and inhibition critically depends on the maturation of GABAergic interneurons during adolescence, suggesting a developmental window of vulnerability for the emergence of psychiatric disorders, including catatonia^28^. Disruptions in the maturation of these interneurons may also contribute to the functional dysconnectivity observed in fMRI studies, particularly involving the medial prefrontal, orbitofrontal, and motor regions in patients with catatonia^19^.

Finally, we observed an increase in the average duration of microstate C in patients with catatonia that was confirmed in the subgroup analysis without benzodiazepines (increases both in duration and time coverage). We did not find any difference in the characteristics of microstate D. Microstate C increases have robustly been replicated across the psychosis spectrum, from ultra-high-risk to first episode psychosis and schizophrenia, and their decrease in patients on treatment suggests a state-dependent process^39^. The difference in microstate C observed in catatonia, compared to non-catatonia individuals, further supports this notion of microstates as potential state markers that reflect gradual changes across disease progression. fMRI studies suggested that microstate C may be related to the default mode network, associated with negative thoughts, mind wandering, self-related thoughts, as well as emotional and interoceptive processing^34^. Several authors consider catatonia to be a primitive fear response, characterized by the onset of stupor in reaction to a perceived threat^78^, because patients experiencing catatonia often report feelings of fear during their episodes^79,80^, and structural alterations in the limbic system have been observed in individuals with catatonia^81^ along with functional changes during the processing of negative emotions^14,82^. An increase in microstate C may thus reflect excessive baseline activation of self-referential and emotional processing networks at the neurophysiological level—processes that are notably disrupted in catatonia.

## LIMITATIONS

Several methodological considerations need to be pointed out. First, the study population was retrospectively selected from a hospital-based real-world health data warehouse. Although each record was reviewed to confirm the presence of catatonic symptoms based on DSM-5 criteria, it is possible that some patients with catatonia were included in the non-catatonia group due to unrecorded or unrecognized catatonia symptoms. As a result, selection bias may have reduced the differences between the two groups. Secondly, patients in the non-catatonia group may exhibit severe psychiatric disorders. Given the naturalistic nature of this clinical dataset, standardized assessments of disorders severity—such as the Positive and Negative Syndrome Scale —were not available. Nonetheless, both groups were comparable in terms of psychotropic medication dosages, including antidepressants and antipsychotics. Moreover, the non-catatonia group enabled us to examine the specific contribution of catatonia on EEG within a realistic clinical framework reflective of routine psychiatric practice, with patients already with schizophrenia or mood disorders. This may also explain why we found relatively small-effect sizes since these markers could already be disrupted in the non-catatonia group. Regarding benzodiazepines, patients with catatonia received higher doses compared to the non-catatonia group, as these medications are the recommended first-line treatment for catatonia^4^. Since psychotropic treatments are known to affect EEG activity^83^, treatment dosages were systematically included as a covariate in the statistical analyses. Additionally, to further control for their influence, a subgroup analysis was conducted on patients who had not received benzodiazepines, in which most of the results were replicated. We also controlled for other potential confounders, including age, sex, and comorbid diagnoses (e.g., neurodevelopmental and neurological disorders). However, residual confounding cannot be completely excluded. From a technical perspective, EEG recordings were acquired during routine clinical practice and were therefore subject to common artifacts. Therefore, we established strict thresholds to remove non-brain signals. Both groups exhibited similar rates of interpolated channels and epochs; however, the ICA rejection rate was slightly higher in the catatonia group. Although it is clear that automatic preprocessing may lead to loss of valuable information, it also allows for the entire EEG analysis pipeline to be fully replicable and ensures its direct applicability on routine EEG. A follow-up project is currently underway employing visual correction to compare automatic and manual preprocessing. Caution is warranted when interpreting gamma-band results, as gamma activity is typically minimal during eyes-closed rest and is more reliably associated with higher-order cognitive processes. Regarding the microstate analysis, it was conducted in the alpha band rather than the broadband, in accordance with the resting-state eyes-closed design of our study^56^. To prevent any bias related to the imbalance between catatonia and non-catatonia group, we avoided building the microstate templates on the whole population. Instead, we built the topomaps on a smaller non-catatonia subgroup before applying them to the rest of the population. Since there was no previous knowledge regarding quantitative EEG markers in catatonia, further prospective longitudinal studies are necessary to replicate and extend our findings.

## CONCLUSION

This study revealed quantitative neurophysiological differences between patients with and without catatonia in a transdiagnostic population with psychotic and mood disorders. Power spectral density, alpha peak, and microstate anomalies in catatonia shed light on its underlying pathophysiology, suggesting a probable neurodevelopmentally-related excitation/inhibition dysregulation. Importantly, our study shows that routine clinical EEG can be leveraged for diagnostic biomarker development, potentially enhancing early detection and treatment.

## Data Availability

All data produced in the present study are available upon reasonable request to the authors

## ACKNOWLEDGEMENTS

Mylene Moyal has received funding from the John Bost Foundation.

This work is part of the TEMPO project (agreement 2024-CER-A-010, PI Anton Iftimovici).

The data warehouse was funded within the framework of France 2030 “Support for the creation of hospital health data warehouses”. This work has also been supported by the French government’s “Investissements d’Avenir” PsyCARE (ANR-18-RHUS-0014) and by a French government grant managed by the Agence Nationale de la Recherche under the France 2030 program, EEG-MIND (ANR-22-EXPR0009).

## CONFLICT OF INTEREST

The Authors have declared that there are no conflicts of interest concerning the subject of this study.

## Notes

### Competing Interest Statement

The authors have declared no competing interest.

### Author Declarations

This study was approved by the GHU Paris Research Ethics Committee on February / 06 / 2024 (agreement number 2024-CER-A-010 TEMPO project).

