## Supplementary Material S2 for "Neurophysiological resting-state EEG markers of catatonia in schizophrenia and mood disorders"

| | | Diagnostic groups | | Tests | |
|---|---|---|---|---|---|
| Demographic | N=621 | Without catatonia (N=519) | Randomly selected non-catatonia patients (N=50) | T or Zscore | p |
| | Age at EEG, M ± SD | 41.7 ± 18.2 | 40.8 ± 18.7 | 11364 | 0.7 |
| | Sex (F/M) | 229 / 290 | 27/23 | -0.28 | 0.77 |
| Diagnostics | Psychotic disorder N(%) | 357(69) | 40(80) | -1.7 | 0.07 |
| | Mood disorder N(%)<br>Bipolar disorder N(%) | 253(48)<br>174(33) | 23(46)<br>17(34) | 0.43<br>-0.07 | 0.6<br>0.94 |
| | Neurodevelopmental disorder N(%) | 39(7) | 4(8) | -0.14 | 0.89 |
| | Neurologic disorder N(%) | 31(6) | 4(8) | -0.5 | 0.61 |
| Treatment | Olanzapine equivalent dosage (mg), M± SD | 32.1 ± 52 | 32.52± 54 | 12257 | 0.59 |
| | Diazepam equivalent dosage (10mg), M± SD | 5.9 ± 30 | 8.7± 31.7 | 11496 | 0.8 |
| | Fluoxetine equivalent dosage (40 mg), M± SD | 0.29 ± 0.65 | 0.53 ± 0.96 | 13393 | **0.03** |

**Table**. Randomly selected non-catatonia patients for microstates maps construction. Abbreviations: Bold font corresponds to significant effects. *Z score proportion test or T test are used for comparison. Results are shown in mean ± standard deviation (M±SD) or absolute number N with corresponding percentages.

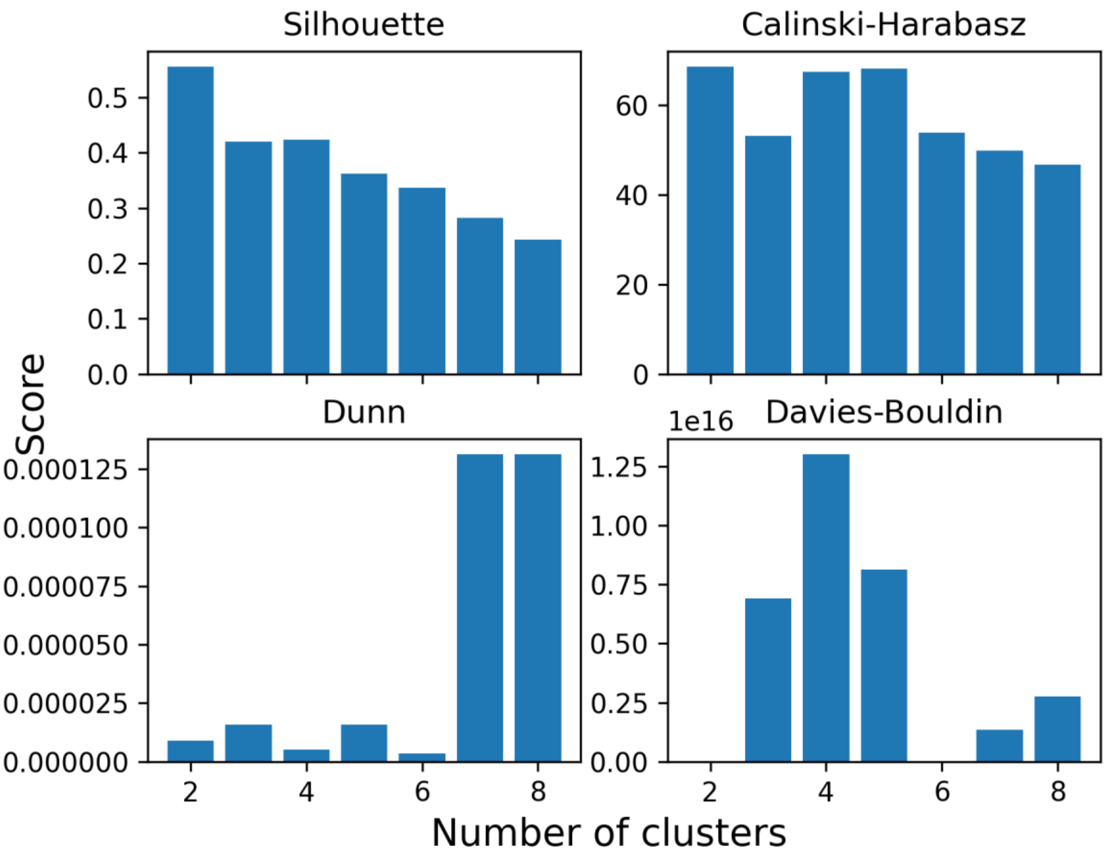


**Figure** the silhouette, Calinski-Harabasz, Davies-Bouldin, and Dunn scores
