## Supplementary Material S8 for "Neurophysiological resting-state EEG markers of catatonia in schizophrenia and mood disorders"

| dependent | names | coef | se | T | pval | CI[2.5%] | CI[97.5%] |
|---|---|---|---|---|---|---|---|
| delta | Intercept | 0.047 | 0.002 | 19.882 | 0 | 0.043 | 0.052 |
| delta | Sexe | -0.001 | 0.002 | -0.831 | 0.407 | -0.005 | 0.002 |
| delta | Agereel | 0 | 0 | -3.462 | 0.001 | 0 | 0 |
| delta | EEG_catatonique | 0.003 | 0.003 | 1.094 | 0.458 | -0.002 | 0.008 |
| delta | total_ola_equivalent | 0 | 0 | 2.112 | 0.035 | 0 | 0 |
| delta | total_fluoxetine_equivalent | -0.001 | 0.001 | -0.659 | 0.51 | -0.003 | 0.002 |
| delta | neurodevelopmental_disorder | 0 | 0.003 | 0.07 | 0.944 | -0.006 | 0.006 |
| delta | neurologic_disorder | 0.007 | 0.003 | 2.161 | 0.031 | 0.001 | 0.013 |
| theta | Intercept | 0.028 | 0.002 | 15.264 | 0 | 0.024 | 0.031 |
| theta | Sexe | -0.001 | 0.001 | -0.88 | 0.38 | -0.004 | 0.001 |
| theta | Agereel | 0 | 0 | -0.22 | 0.826 | 0 | 0 |
| theta | EEG_catatonique | 0.001 | 0.002 | 0.556 | 0.579 | -0.003 | 0.005 |
| theta | total_ola_equivalent | 0 | 0 | 0.435 | 0.664 | 0 | 0 |
| theta | total_fluoxetine_equivalent | 0 | 0.001 | 0.143 | 0.886 | -0.002 | 0.002 |
| theta | neurodevelopmental_disorder | -0.003 | 0.002 | -1.081 | 0.281 | -0.007 | 0.002 |
| theta | neurologic_disorder | 0.005 | 0.002 | 2.177 | 0.03 | 0.001 | 0.01 |
| alpha | Intercept | 0.037 | 0.002 | 19.702 | 0 | 0.033 | 0.04 |
| alpha | Sexe | -0.001 | 0.001 | -0.429 | 0.668 | -0.003 | 0.002 |
| alpha | Agereel | 0 | 0 | -2 | 0.046 | 0 | 0 |
| alpha | EEG_catatonique | -0.004 | 0.002 | -2.113 | 0.088 | -0.008 | 0 |
| alpha | total_ola_equivalent | 0 | 0 | -1.718 | 0.087 | 0 | 0 |
| alpha | total_fluoxetine_equivalent | -0.002 | 0.001 | -1.72 | 0.086 | -0.004 | 0 |
| alpha | neurodevelopmental_disorder | 0.003 | 0.002 | 1.131 | 0.259 | -0.002 | 0.008 |
| alpha | neurologic_disorder | -0.002 | 0.002 | -0.789 | 0.431 | -0.007 | 0.003 |
| beta | Intercept | 0.005 | 0 | 11.28 | 0 | 0.004 | 0.006 |
| beta | Sexe | 0 | 0 | 1.216 | 0.225 | 0 | 0.001 |
| beta | Agereel | 0 | 0 | 4.486 | 0 | 0 | 0 |
| beta | EEG_catatonique | 0 | 0 | -0.571 | 0.579 | -0.001 | 0.001 |
| beta | total_ola_equivalent | 0 | 0 | -0.986 | 0.325 | 0 | 0 |
| beta | total_fluoxetine_equivalent | 0 | 0 | 1.309 | 0.191 | 0 | 0.001 |
| beta | neurodevelopmental_disorder | 0 | 0.001 | -0.18 | 0.857 | -0.001 | 0.001 |
| beta | neurologic_disorder | -0.002 | 0.001 | -2.965 | 0.003 | -0.003 | -0.001 |

| | | | | | | | |
|---|---|---|---|---|---|---|---|
| gamma | Intercept | 0.001 | 0 | 5.299 | 0 | 0.001 | 0.001 |
| gamma | Sexe | 0 | 0 | 2.274 | 0.024 | 0 | 0.001 |
| gamma | Agereel | 0 | 0 | 2.444 | 0.015 | 0 | 0 |
| gamma | EEG_catatonique | 0.001 | 0 | 3.08 | 0.011 | 0 | 0.001 |
| gamma | total_ola_equivalent | 0 | 0 | 0.787 | 0.432 | 0 | 0 |
| gamma | total_fluoxetine_equivalent | 0 | 0 | 2.363 | 0.019 | 0 | 0 |
| gamma | neurodevelopmental_disorder | 0 | 0 | 0.113 | 0.91 | 0 | 0 |
| gamma | neurologic_disorder | 0 | 0 | -0.907 | 0.365 | -0.001 | 0 |
| alpha_peak | Intercept | 9.904 | 0.122 | 81.305 | 0 | 9.664 | 10.144 |
| alpha_peak | Sexe | 0.098 | 0.091 | 1.069 | 0.286 | -0.082 | 0.278 |
| alpha_peak | Agereel | -0.01 | 0.002 | -4.202 | 0 | -0.015 | -0.005 |
| alpha_peak | EEG_catatonique | -0.317 | 0.129 | -2.461 | 0.014 | -0.571 | -0.064 |
| alpha_peak | total_ola_equivalent | -0.001 | 0.001 | -1.036 | 0.301 | -0.003 | 0.001 |
| alpha_peak | total_fluoxetine_equivalent | 0.081 | 0.067 | 1.209 | 0.227 | -0.051 | 0.214 |
| alpha_peak | neurodevelopmental_disorder | 0.303 | 0.161 | 1.881 | 0.061 | -0.014 | 0.621 |
| alpha_peak | neurologic_disorder | -0.185 | 0.162 | -1.142 | 0.254 | -0.504 | 0.134 |
| C_occurrences | Intercept | 3.224 | 0.287 | 11.22 | 0 | 2.659 | 3.789 |
| C_occurrences | Sexe | -0.15 | 0.212 | -0.711 | 0.478 | -0.567 | 0.266 |
| C_occurrences | Agereel | -0.006 | 0.006 | -1.066 | 0.287 | -0.017 | 0.005 |
| C_occurrences | EEG_catatonique | 0.38 | 0.289 | 1.317 | 0.189 | -0.188 | 0.948 |
| C_occurrences | total_ola_equivalent | -0.002 | 0.003 | -0.735 | 0.463 | -0.007 | 0.003 |
| C_occurrences | total_fluoxetine_equivalent | -0.061 | 0.165 | -0.368 | 0.713 | -0.386 | 0.264 |
| C_occurrences | neurodevelopmental_disorder | 0.072 | 0.366 | 0.198 | 0.843 | -0.648 | 0.793 |
| C_occurrences | neurologic_disorder | 0.083 | 0.372 | 0.223 | 0.824 | -0.649 | 0.814 |
| C_timecov | Intercept | 0.155 | 0.018 | 8.671 | 0 | 0.12 | 0.19 |
| C_timecov | Sexe | -0.001 | 0.013 | -0.081 | 0.936 | -0.027 | 0.025 |
| C_timecov | Agereel | 0 | 0 | -1.215 | 0.225 | -0.001 | 0 |
| C_timecov | EEG_catatonique | 0.035 | 0.018 | 1.973 | 0.049 | 0 | 0.071 |

| | | | | | | | |
|---|---|---|---|---|---|---|---|
| C_timecov | total_ola_equivalent | 0 | 0 | -0.323 | 0.747 | 0 | 0 |
| C_timecov | total_fluoxetine_equivalent | -0.008 | 0.01 | -0.743 | 0.458 | -0.028 | 0.013 |
| C_timecov | neurodevelopmental_disorder | -0.009 | 0.023 | -0.383 | 0.702 | -0.054 | 0.036 |
| C_timecov | neurologic_disorder | 0.001 | 0.023 | 0.035 | 0.972 | -0.045 | 0.046 |
| C_meandurs | Intercept | 0.043 | 0.002 | 27.707 | 0 | 0.04 | 0.046 |
| C_meandurs | Sexe | 0.001 | 0.001 | 0.481 | 0.631 | -0.002 | 0.003 |
| C_meandurs | Agereel | 0 | 0 | -1.432 | 0.153 | 0 | 0 |
| C_meandurs | EEG_catatonique | 0.005 | 0.002 | 2.991 | 0.003 | 0.002 | 0.008 |
| C_meandurs | total_ola_equivalent | 0 | 0 | 0.165 | 0.869 | 0 | 0 |
| C_meandurs | total_fluoxetine_equivalent | -0.001 | 0.001 | -0.662 | 0.508 | -0.002 | 0.001 |
| C_meandurs | neurodevelopmental_disorder | -0.001 | 0.002 | -0.682 | 0.496 | -0.005 | 0.003 |
| C_meandurs | neurologic_disorder | 0 | 0.002 | 0.172 | 0.863 | -0.004 | 0.004 |
| D_occurrences | Intercept | 4.107 | 0.291 | 14.096 | 0 | 3.534 | 4.68 |
| D_occurrences | Sexe | 0.231 | 0.215 | 1.075 | 0.283 | -0.192 | 0.653 |
| D_occurrences | Agereel | -0.007 | 0.006 | -1.159 | 0.247 | -0.018 | 0.005 |
| D_occurrences | EEG_catatonique | -0.012 | 0.293 | -0.042 | 0.967 | -0.588 | 0.564 |
| D_occurrences | total_ola_equivalent | 0 | 0.003 | 0.053 | 0.958 | -0.005 | 0.006 |
| D_occurrences | total_fluoxetine_equivalent | -0.116 | 0.168 | -0.694 | 0.489 | -0.446 | 0.214 |
| D_occurrences | neurodevelopmental_disorder | 0.063 | 0.371 | 0.171 | 0.864 | -0.667 | 0.794 |
| D_occurrences | neurologic_disorder | -0.069 | 0.377 | -0.184 | 0.854 | -0.811 | 0.672 |
| D_timecov | Intercept | 0.261 | 0.033 | 7.812 | 0 | 0.196 | 0.327 |
| D_timecov | Sexe | 0.036 | 0.025 | 1.476 | 0.141 | -0.012 | 0.085 |
| D_timecov | Agereel | -0.001 | 0.001 | -0.91 | 0.363 | -0.002 | 0.001 |
| D_timecov | EEG_catatonique | 0.001 | 0.034 | 0.023 | 0.982 | -0.065 | 0.067 |
| D_timecov | total_ola_equivalent | 0 | 0 | -0.23 | 0.818 | -0.001 | 0.001 |
| D_timecov | total_fluoxetine_equivalent | 0.003 | 0.019 | 0.147 | 0.883 | -0.035 | 0.041 |

| | | | | | | | |
|---|---|---|---|---|---|---|---|
| D_timecov | neurodevelopmental_disorder | -0.038 | 0.043 | -0.88 | 0.379 | -0.121 | 0.046 |
| D_timecov | neurologic_disorder | 0.047 | 0.043 | 1.087 | 0.278 | -0.038 | 0.132 |
| D_meandurs | Intercept | 0.058 | 0.007 | 8.571 | 0 | 0.045 | 0.071 |
| D_meandurs | Sexe | 0.006 | 0.005 | 1.161 | 0.246 | -0.004 | 0.016 |
| D_meandurs | Agereel | 0 | 0 | -0.355 | 0.723 | 0 | 0 |
| D_meandurs | EEG_catatonique | -0.001 | 0.007 | -0.078 | 0.938 | -0.014 | 0.013 |
| D_meandurs | total_ola_equivalent | 0 | 0 | -0.39 | 0.697 | 0 | 0 |
| D_meandurs | total_fluoxetine_equivalent | 0.001 | 0.004 | 0.283 | 0.777 | -0.007 | 0.009 |
| D_meandurs | neurodevelopmental_disorder | -0.006 | 0.009 | -0.719 | 0.472 | -0.023 | 0.011 |
| D_meandurs | neurologic_disorder | 0.017 | 0.009 | 1.983 | 0.048 | 0 | 0.035 |

Table Results of linear regression model in the subgroup without benzodiazepines between EEG features and catatonia status with covariates of age, sex, treatment (total equivalent of diazepam, olanzapine and fluoxetine), associated diagnosis (neurodevelopmental or neurological disorder). Abbreviations: Bold font corresponds to significant effects. Coef: coefficient; SD: standard deviation; CI[2.5%] CI[97.5%] to the confidence intervals at 2.5 and 97.5% and pval for p.value.
