## Supplementary Material S7 for "Neurophysiological resting-state EEG markers of catatonia in schizophrenia and mood disorders"

| dependent | names | coef | se | T | pval | CI[2.5 %] | CI[97.5 %] |
|---|---|---|---|---|---|---|---|
| delta | Intercept | 0.044 | 0.004 | 10.192 | 0 | 0.035 | 0.052 |
| delta | Sexe | -0.003 | 0.003 | -0.995 | 0.321 | -0.008 | 0.003 |
| delta | Agereel | 0 | 0 | -1.533 | 0.127 | 0 | 0 |
| delta | EEG_catatonique | 0.003 | 0.003 | 0.955 | 0.341 | -0.004 | 0.01 |
| delta | total_diazepam_equivalent | 0 | 0 | 0.676 | 0.5 | 0 | 0 |
| delta | total_ola_equivalent | 0 | 0 | -0.441 | 0.659 | 0 | 0 |
| delta | total_fluoxetine_equivalent | 0 | 0.002 | -0.258 | 0.796 | -0.004 | 0.003 |
| delta | neurodevelopmental_disorder | -0.005 | 0.007 | -0.755 | 0.451 | -0.018 | 0.008 |
| delta | neurologic_disorder | 0.003 | 0.004 | 0.831 | 0.407 | -0.005 | 0.011 |
| theta | Intercept | 0.02 | 0.003 | 6.581 | 0 | 0.014 | 0.026 |
| theta | Sexe | -0.001 | 0.002 | -0.325 | 0.745 | -0.004 | 0.003 |
| theta | Agereel | 0 | 0 | 2.128 | 0.035 | 0 | 0 |
| theta | EEG_catatonique | -0.002 | 0.002 | -0.956 | 0.341 | -0.007 | 0.002 |
| theta | total_diazepam_equivalent | 0 | 0 | -0.334 | 0.739 | 0 | 0 |
| theta | total_ola_equivalent | 0 | 0 | 0.557 | 0.578 | 0 | 0 |
| theta | total_fluoxetine_equivalent | 0 | 0.001 | 0.094 | 0.925 | -0.002 | 0.002 |
| theta | neurodevelopmental_disorder | -0.008 | 0.005 | -1.631 | 0.105 | -0.017 | 0.002 |
| theta | neurologic_disorder | 0.007 | 0.003 | 2.622 | 0.01 | 0.002 | 0.013 |
| alpha | Intercept | 0.035 | 0.003 | 11.16 | 0 | 0.029 | 0.041 |
| alpha | Sexe | 0.001 | 0.002 | 0.298 | 0.766 | -0.003 | 0.004 |
| alpha | Agereel | 0 | 0 | -1.437 | 0.152 | 0 | 0 |
| alpha | EEG_catatonique | -0.005 | 0.002 | -1.9 | 0.289 | -0.01 | 0 |
| alpha | total_diazepam_equivalent | 0 | 0 | 0.219 | 0.827 | 0 | 0 |
| alpha | total_ola_equivalent | 0 | 0 | 0.144 | 0.886 | 0 | 0 |
| alpha | total_fluoxetine_equivalent | 0 | 0.001 | 0.34 | 0.734 | -0.002 | 0.003 |
| alpha | neurodevelopmental_disorder | 0 | 0.005 | 0.006 | 0.995 | -0.01 | 0.01 |
| alpha | neurologic_disorder | -0.003 | 0.003 | -0.856 | 0.393 | -0.008 | 0.003 |
| beta | Intercept | 0.007 | 0.001 | 8.738 | 0 | 0.006 | 0.009 |
| beta | Sexe | 0 | 0.001 | 0.811 | 0.418 | -0.001 | 0.001 |
| beta | Agereel | 0 | 0 | 0.285 | 0.776 | 0 | 0 |
| beta | EEG_catatonique | 0.001 | 0.001 | 1.012 | 0.341 | -0.001 | 0.002 |
| beta | total_diazepam_equivalent | 0 | 0 | -0.423 | 0.673 | 0 | 0 |

| | | | | | | | |
|---|---|---|---|---|---|---|---|
| beta | total_ola_equivalent | 0 | 0 | 0.147 | 0.883 | 0 | 0 |
| beta | total_fluoxetine_equivalent | 0 | 0 | -0.376 | 0.708 | -0.001 | 0.001 |
| beta | neurodevelopmental_disorder | 0.002 | 0.001 | 1.304 | 0.194 | -0.001 | 0.004 |
| beta | neurologic_disorder | -0.001 | 0.001 | -1.873 | 0.063 | -0.003 | 0 |
| gamma | Intercept | 0.001 | 0 | 4.023 | 0 | 0.001 | 0.002 |
| gamma | Sexe | 0 | 0 | 0.181 | 0.857 | 0 | 0 |
| gamma | Agereel | 0 | 0 | 1.555 | 0.122 | 0 | 0 |
| gamma | EEG_catatonique | 0 | 0 | 1.582 | 0.289 | 0 | 0.001 |
| gamma | total_diazepam_equivalent | 0 | 0 | -0.213 | 0.831 | 0 | 0 |
| gamma | total_ola_equivalent | 0 | 0 | -1.042 | 0.299 | 0 | 0 |
| gamma | total_fluoxetine_equivalent | 0 | 0 | 0.742 | 0.459 | 0 | 0 |
| gamma | neurodevelopmental_disorder | 0.001 | 0.001 | 2.025 | 0.044 | 0 | 0.002 |
| gamma | neurologic_disorder | 0 | 0 | -0.763 | 0.447 | -0.001 | 0 |
| alpha_peak | Intercept | 10.099 | 0.201 | 50.349 | 0 | 9.704 | 10.495 |
| alpha_peak | Sexe | 0.053 | 0.125 | 0.42 | 0.675 | -0.194 | 0.299 |
| alpha_peak | Agereel | -0.013 | 0.003 | -4.113 | 0 | -0.019 | -0.007 |
| alpha_peak | EEG_catatonique | -0.111 | 0.161 | -0.689 | 0.491 | -0.428 | 0.206 |
| alpha_peak | total_diazepam_equivalent | -0.001 | 0.001 | -0.373 | 0.709 | -0.003 | 0.002 |
| alpha_peak | total_ola_equivalent | -0.001 | 0.002 | -0.325 | 0.746 | -0.005 | 0.004 |
| alpha_peak | total_fluoxetine_equivalent | 0.018 | 0.078 | 0.235 | 0.814 | -0.136 | 0.173 |
| alpha_peak | neurodevelopmental_disorder | 0.374 | 0.312 | 1.199 | 0.232 | -0.242 | 0.991 |
| alpha_peak | neurologic_disorder | -0.317 | 0.189 | -1.676 | 0.096 | -0.69 | 0.056 |
| C_occurrences | Intercept | 3.189 | 0.429 | 7.431 | 0 | 2.342 | 4.037 |
| C_occurrences | Sexe | -0.152 | 0.271 | -0.562 | 0.575 | -0.687 | 0.383 |
| C_occurrences | Agereel | -0.004 | 0.007 | -0.555 | 0.58 | -0.018 | 0.01 |
| C_occurrences | EEG_catatonique | -0.049 | 0.342 | -0.143 | 0.887 | -0.723 | 0.626 |
| C_occurrences | total_ola_equivalent | 0 | 0.005 | 0.067 | 0.947 | -0.01 | 0.01 |
| C_occurrences | total_fluoxetine_equivalent | -0.079 | 0.188 | -0.421 | 0.674 | -0.451 | 0.292 |
| C_occurrences | total_diazepam_equivalent | 0.005 | 0.003 | 1.537 | 0.126 | -0.001 | 0.011 |

| | | | | | | | |
|---|---|---|---|---|---|---|---|
| C_occurrences | neurodevelopmental_disorder | -0.274 | 0.661 | -0.415 | 0.679 | -1.58 | 1.031 |
| C_occurrences | neurologic_disorder | -0.16 | 0.4 | -0.4 | 0.69 | -0.949 | 0.629 |
| C_timecov | Intercept | 0.132 | 0.024 | 5.413 | 0 | 0.084 | 0.18 |
| C_timecov | Sexe | -0.003 | 0.015 | -0.17 | 0.865 | -0.033 | 0.028 |
| C_timecov | Agereel | 0 | 0 | -0.013 | 0.989 | -0.001 | 0.001 |
| C_timecov | EEG_catatonique | -0.004 | 0.019 | -0.218 | 0.828 | -0.043 | 0.034 |
| C_timecov | total_ola_equivalent | 0 | 0 | 0.285 | 0.776 | 0 | 0.001 |
| C_timecov | total_fluoxetine_equivalent | -0.005 | 0.011 | -0.455 | 0.649 | -0.026 | 0.016 |
| C_timecov | total_diazepam_equivalent | 0 | 0 | 2.038 | 0.043 | 0 | 0.001 |
| C_timecov | neurodevelopmental_disorder | -0.018 | 0.038 | -0.466 | 0.642 | -0.092 | 0.057 |
| C_timecov | neurologic_disorder | -0.013 | 0.023 | -0.57 | 0.569 | -0.058 | 0.032 |
| C_meandurs | Intercept | 0.04 | 0.002 | 19.064 | 0 | 0.036 | 0.044 |
| C_meandurs | Sexe | 0 | 0.001 | 0.337 | 0.736 | -0.002 | 0.003 |
| C_meandurs | Agereel | 0 | 0 | 0.671 | 0.503 | 0 | 0 |
| C_meandurs | EEG_catatonique | 0 | 0.002 | -0.031 | 0.975 | -0.003 | 0.003 |
| C_meandurs | total_ola_equivalent | 0 | 0 | 0.055 | 0.957 | 0 | 0 |
| C_meandurs | total_fluoxetine_equivalent | 0 | 0.001 | 0.074 | 0.941 | -0.002 | 0.002 |
| C_meandurs | total_diazepam_equivalent | 0 | 0 | 1.865 | 0.064 | 0 | 0 |
| C_meandurs | neurodevelopmental_disorder | -0.001 | 0.003 | -0.35 | 0.727 | -0.007 | 0.005 |
| C_meandurs | neurologic_disorder | -0.001 | 0.002 | -0.434 | 0.665 | -0.005 | 0.003 |
| D_occurrences | Intercept | 4.484 | 0.454 | 9.87 | 0 | 3.587 | 5.381 |
| D_occurrences | Sexe | 0.184 | 0.287 | 0.642 | 0.522 | -0.382 | 0.75 |
| D_occurrences | Agereel | -0.013 | 0.007 | -1.785 | 0.076 | -0.028 | 0.001 |
| D_occurrences | EEG_catatonique | -0.052 | 0.362 | -0.144 | 0.886 | -0.766 | 0.662 |
| D_occurrences | total_ola_equivalent | 0.002 | 0.005 | 0.426 | 0.67 | -0.008 | 0.013 |
| D_occurrences | total_fluoxetine_equivalent | -0.152 | 0.199 | -0.765 | 0.445 | -0.545 | 0.241 |
| D_occurrences | total_diazepam_equivalent | 0 | 0.003 | 0.081 | 0.936 | -0.006 | 0.007 |
| D_occurrences | neurodevelopmental_disorder | -0.57 | 0.7 | -0.814 | 0.417 | -1.952 | 0.812 |

| | | | | | | | |
|---|---|---|---|---|---|---|---|
| D_occurrences | neurologic_disorder | 0.09 | 0.423 | 0.213 | 0.831 | -0.745 | 0.926 |
| D_timecov | Intercept | 0.233 | 0.052 | 4.482 | 0 | 0.131 | 0.336 |
| D_timecov | Sexe | 0.027 | 0.033 | 0.819 | 0.414 | -0.038 | 0.092 |
| D_timecov | Agereel | 0 | 0.001 | -0.248 | 0.805 | -0.002 | 0.001 |
| D_timecov | EEG_catatonique | -0.063 | 0.041 | -1.521 | 0.13 | -0.145 | 0.019 |
| D_timecov | total_ola_equivalent | 0.001 | 0.001 | 0.842 | 0.401 | -0.001 | 0.002 |
| D_timecov | total_fluoxetine_equivalent | 0.02 | 0.023 | 0.885 | 0.377 | -0.025 | 0.065 |
| D_timecov | total_diazepam_equivalent | 0 | 0 | -0.482 | 0.63 | -0.001 | 0.001 |
| D_timecov | neurodevelopmental_disorder | -0.043 | 0.08 | -0.532 | 0.595 | -0.201 | 0.116 |
| D_timecov | neurologic_disorder | 0.054 | 0.048 | 1.109 | 0.269 | -0.042 | 0.15 |
| D_meandurs | Intercept | 0.047 | 0.01 | 4.634 | 0 | 0.027 | 0.068 |
| D_meandurs | Sexe | 0.007 | 0.006 | 1.02 | 0.309 | -0.006 | 0.019 |
| D_meandurs | Agereel | 0 | 0 | 0.518 | 0.605 | 0 | 0 |
| D_meandurs | EEG_catatonique | -0.013 | 0.008 | -1.644 | 0.102 | -0.029 | 0.003 |
| D_meandurs | total_ola_equivalent | 0 | 0 | 0.709 | 0.48 | 0 | 0 |
| D_meandurs | total_fluoxetine_equivalent | 0.005 | 0.004 | 1.157 | 0.249 | -0.004 | 0.014 |
| D_meandurs | total_diazepam_equivalent | 0 | 0 | -0.512 | 0.609 | 0 | 0 |
| D_meandurs | neurodevelopmental_disorder | -0.007 | 0.016 | -0.426 | 0.671 | -0.038 | 0.024 |
| D_meandurs | neurologic_disorder | 0.016 | 0.01 | 1.661 | 0.099 | -0.003 | 0.035 |

**Table** Results of linear regression model in the mood subgroup between EEG features and catatonia status with covariates of age, sex, treatment (total equivalent of diazepam, olanzapine and fluoxetine), associated diagnosis (neurodevelopmental or neurological disorder). Abbreviations: Bold font corresponds to significant effects. Coef: coefficient; SD: standard deviation; CI[2.5%] CI[97.5%] to the confidence intervals at 2.5 and 97.5% and pval for p.value.
