## Supplementary Material S6 for "Neurophysiological resting-state EEG markers of catatonia in schizophrenia and mood disorders"

| dependent | names | coef | se | T | pval | CI[2.5 %] | CI[97.5 %] |
|---|---|---|---|---|---|---|---|
| delta | Intercept | 0.046 | 0.002 | 18.615 | 0 | 0.041 | 0.05 |
| delta | Sexe | -0.002 | 0.002 | -0.959 | 0.338 | -0.005 | 0.002 |
| delta | Agereel | 0 | 0 | -2.394 | 0.017 | 0 | 0 |
| delta | EEG_catatonique | 0.004 | 0.002 | 1.705 | 0.148 | -0.001 | 0.009 |
| delta | total_diazepam_equivalent | 0 | 0 | -1.042 | 0.298 | 0 | 0 |
| delta | total_ola_equivalent | 0 | 0 | 1.64 | 0.102 | 0 | 0 |
| delta | total_fluoxetine_equivalent | -0.002 | 0.002 | -1.412 | 0.159 | -0.005 | 0.001 |
| delta | neurodevelopmental_disorder | 0.004 | 0.003 | 1.461 | 0.145 | -0.002 | 0.01 |
| delta | neurologic_disorder | 0.011 | 0.004 | 2.961 | 0.003 | 0.004 | 0.018 |
| theta | Intercept | 0.025 | 0.002 | 14.463 | 0 | 0.022 | 0.029 |
| theta | Sexe | -0.002 | 0.001 | -1.214 | 0.225 | -0.004 | 0.001 |
| theta | Agereel | 0 | 0 | 1.065 | 0.287 | 0 | 0 |
| theta | EEG_catatonique | 0.002 | 0.002 | 0.953 | 0.426 | -0.002 | 0.005 |
| theta | total_diazepam_equivalent | 0 | 0 | -0.437 | 0.662 | 0 | 0 |
| theta | total_ola_equivalent | 0 | 0 | 0.363 | 0.717 | 0 | 0 |
| theta | total_fluoxetine_equivalent | 0.001 | 0.001 | 0.564 | 0.573 | -0.001 | 0.003 |
| theta | neurodevelopmental_disorder | -0.001 | 0.002 | -0.348 | 0.728 | -0.005 | 0.003 |
| theta | neurologic_disorder | 0.005 | 0.003 | 1.743 | 0.082 | -0.001 | 0.01 |
| alpha | Intercept | 0.036 | 0.002 | 20.31 | 0 | 0.033 | 0.04 |
| alpha | Sexe | -0.001 | 0.001 | -0.533 | 0.594 | -0.003 | 0.002 |
| alpha | Agereel | 0 | 0 | -2.162 | 0.031 | 0 | 0 |
| alpha | EEG_catatonique | -0.005 | 0.002 | -2.63 | 0.044 | -0.008 | -0.001 |
| alpha | total_diazepam_equivalent | 0 | 0 | 1.656 | 0.098 | 0 | 0 |
| alpha | total_ola_equivalent | 0 | 0 | -3.71 | 0 | 0 | 0 |
| alpha | total_fluoxetine_equivalent | 0 | 0.001 | 0.169 | 0.866 | -0.002 | 0.002 |
| alpha | neurodevelopmental_disorder | 0 | 0.002 | 0.157 | 0.875 | -0.004 | 0.005 |
| alpha | neurologic_disorder | -0.004 | 0.003 | -1.347 | 0.179 | -0.009 | 0.002 |
| beta | Intercept | 0.006 | 0 | 11.576 | 0 | 0.005 | 0.007 |
| beta | Sexe | 0 | 0 | 1.143 | 0.254 | 0 | 0.001 |
| beta | Agereel | 0 | 0 | 2.412 | 0.016 | 0 | 0 |
| beta | EEG_catatonique | 0 | 0.001 | -0.543 | 0.588 | -0.001 | 0.001 |
| beta | total_diazepam_equivalent | 0 | 0 | 0.284 | 0.777 | 0 | 0 |

| | | | | | | | |
|---|---|---|---|---|---|---|---|
| beta | total_ola_equivalent | 0 | 0 | 1.051 | 0.294 | 0 | 0 |
| beta | total_fluoxetine_equivalent | 0 | 0 | 0.296 | 0.768 | -0.001 | 0.001 |
| beta | neurodevelopmental_disorder | 0 | 0.001 | -0.794 | 0.428 | -0.002 | 0.001 |
| beta | neurologic_disorder | -0.002 | 0.001 | -2.634 | 0.009 | -0.003 | 0 |
| gamma | Intercept | 0.001 | 0 | 6.933 | 0 | 0.001 | 0.001 |
| gamma | Sexe | 0 | 0 | 4.069 | 0 | 0 | 0.001 |
| gamma | Agereel | 0 | 0 | 1.886 | 0.06 | 0 | 0 |
| gamma | EEG_catatonique | 0 | 0 | 1.974 | 0.123 | 0 | 0.001 |
| gamma | total_diazepam_equivalent | 0 | 0 | -1.625 | 0.105 | 0 | 0 |
| gamma | total_ola_equivalent | 0 | 0 | 1.153 | 0.249 | 0 | 0 |
| gamma | total_fluoxetine_equivalent | 0 | 0 | 1.169 | 0.243 | 0 | 0 |
| gamma | neurodevelopmental_disorder | 0 | 0 | -1.065 | 0.287 | -0.001 | 0 |
| gamma | neurologic_disorder | 0 | 0 | -0.397 | 0.692 | -0.001 | 0 |
| alpha_peak | Intercept | 9.805 | 0.123 | 79.701 | 0 | 9.563 | 10.047 |
| alpha_peak | Sexe | 0.146 | 0.092 | 1.583 | 0.114 | -0.035 | 0.326 |
| alpha_peak | Agereel | -0.01 | 0.003 | -3.384 | 0.001 | -0.016 | -0.004 |
| alpha_peak | EEG_catatonique | -0.324 | 0.126 | -2.578 | 0.01 | -0.57 | -0.077 |
| alpha_peak | total_diazepam_equivalent | -0.001 | 0.002 | -0.556 | 0.578 | -0.005 | 0.003 |
| alpha_peak | total_ola_equivalent | 0 | 0.001 | 0.355 | 0.723 | -0.001 | 0.002 |
| alpha_peak | total_fluoxetine_equivalent | 0.058 | 0.076 | 0.763 | 0.446 | -0.091 | 0.207 |
| alpha_peak | neurodevelopmental_disorder | 0.195 | 0.152 | 1.28 | 0.201 | -0.104 | 0.494 |
| alpha_peak | neurologic_disorder | -0.293 | 0.184 | -1.592 | 0.112 | -0.655 | 0.069 |
| C_occurrences | Intercept | 2.91 | 0.282 | 10.305 | 0 | 2.355 | 3.465 |
| C_occurrences | Sexe | -0.145 | 0.208 | -0.697 | 0.486 | -0.554 | 0.264 |
| C_occurrences | Agereel | 0.001 | 0.007 | 0.197 | 0.844 | -0.012 | 0.014 |
| C_occurrences | EEG_catatonique | 0.019 | 0.276 | 0.069 | 0.945 | -0.524 | 0.562 |
| C_occurrences | total_ola_equivalent | -0.002 | 0.002 | -0.96 | 0.338 | -0.005 | 0.002 |
| C_occurrences | total_fluoxetine_equivalent | 0.219 | 0.178 | 1.235 | 0.218 | -0.13 | 0.569 |
| C_occurrences | total_diazepam_equivalent | -0.001 | 0.004 | -0.269 | 0.788 | -0.009 | 0.007 |

| | | | | | | | |
|---|---|---|---|---|---|---|---|
| C_occurrences | neurodevelopmental_disorder | 0.281 | 0.349 | 0.807 | 0.42 | -0.404 | 0.967 |
| C_occurrences | neurologic_disorder | 0.176 | 0.43 | 0.41 | 0.682 | -0.67 | 1.023 |
| C_timecov | Intercept | 0.135 | 0.017 | 7.875 | 0 | 0.101 | 0.169 |
| C_timecov | Sexe | -0.003 | 0.013 | -0.246 | 0.806 | -0.028 | 0.022 |
| C_timecov | Agereel | 0 | 0 | 0.069 | 0.945 | -0.001 | 0.001 |
| C_timecov | EEG_catatonique | 0.005 | 0.017 | 0.302 | 0.762 | -0.028 | 0.038 |
| C_timecov | total_ola_equivalent | 0 | 0 | -0.66 | 0.51 | 0 | 0 |
| C_timecov | total_fluoxetine_equivalent | 0.019 | 0.011 | 1.791 | 0.074 | -0.002 | 0.041 |
| C_timecov | total_diazepam_equivalent | 0 | 0 | -0.651 | 0.515 | -0.001 | 0 |
| C_timecov | neurodevelopmental_disorder | 0.006 | 0.021 | 0.273 | 0.785 | -0.036 | 0.047 |
| C_timecov | neurologic_disorder | 0.011 | 0.026 | 0.427 | 0.67 | -0.04 | 0.063 |
| C_meandurs | Intercept | 0.042 | 0.002 | 27.392 | 0 | 0.039 | 0.045 |
| C_meandurs | Sexe | 0 | 0.001 | 0.023 | 0.981 | -0.002 | 0.002 |
| C_meandurs | Agereel | 0 | 0 | -0.353 | 0.724 | 0 | 0 |
| C_meandurs | EEG_catatonique | 0.002 | 0.002 | 1.169 | 0.243 | -0.001 | 0.005 |
| C_meandurs | total_ola_equivalent | 0 | 0 | -0.548 | 0.584 | 0 | 0 |
| C_meandurs | total_fluoxetine_equivalent | 0.001 | 0.001 | 1.473 | 0.142 | 0 | 0.003 |
| C_meandurs | total_diazepam_equivalent | 0 | 0 | -0.953 | 0.341 | 0 | 0 |
| C_meandurs | neurodevelopmental_disorder | -0.001 | 0.002 | -0.706 | 0.481 | -0.005 | 0.002 |
| C_meandurs | neurologic_disorder | 0.001 | 0.002 | 0.593 | 0.554 | -0.003 | 0.006 |
| D_occurrences | Intercept | 4.076 | 0.266 | 15.347 | 0 | 3.554 | 4.598 |
| D_occurrences | Sexe | 0.223 | 0.196 | 1.137 | 0.256 | -0.162 | 0.607 |
| D_occurrences | Agereel | -0.002 | 0.006 | -0.243 | 0.808 | -0.014 | 0.011 |
| D_occurrences | EEG_catatonique | -0.079 | 0.26 | -0.304 | 0.762 | -0.589 | 0.432 |
| D_occurrences | total_ola_equivalent | -0.001 | 0.002 | -0.744 | 0.457 | -0.004 | 0.002 |
| D_occurrences | total_fluoxetine_equivalent | -0.261 | 0.167 | -1.565 | 0.118 | -0.59 | 0.067 |
| D_occurrences | total_diazepam_equivalent | -0.005 | 0.004 | -1.279 | 0.202 | -0.013 | 0.003 |
| D_occurrences | neurodevelopmental_disorder | 0.081 | 0.328 | 0.248 | 0.804 | -0.564 | 0.727 |

| | | | | | | | |
|---|---|---|---|---|---|---|---|
| D_occurrences | neurologic_disorder | -0.465 | 0.405 | -1.148 | 0.252 | -1.261 | 0.331 |
| D_timecov | Intercept | 0.267 | 0.032 | 8.317 | 0 | 0.204 | 0.33 |
| D_timecov | Sexe | 0.008 | 0.024 | 0.346 | 0.73 | -0.038 | 0.055 |
| D_timecov | Agereel | 0 | 0.001 | -0.099 | 0.921 | -0.002 | 0.001 |
| D_timecov | EEG_catatonique | 0.012 | 0.031 | 0.388 | 0.698 | -0.05 | 0.074 |
| D_timecov | total_ola_equivalent | 0 | 0 | -0.493 | 0.622 | 0 | 0 |
| D_timecov | total_fluoxetine_equivalent | -0.006 | 0.02 | -0.286 | 0.775 | -0.045 | 0.034 |
| D_timecov | total_diazepam_equivalent | -0.001 | 0 | -1.122 | 0.262 | -0.001 | 0 |
| D_timecov | neurodevelopmental_disorder | -0.022 | 0.04 | -0.548 | 0.584 | -0.1 | 0.056 |
| D_timecov | neurologic_disorder | -0.042 | 0.049 | -0.867 | 0.387 | -0.139 | 0.054 |
| D_meandurs | Intercept | 0.062 | 0.007 | 8.561 | 0 | 0.048 | 0.076 |
| D_meandurs | Sexe | -0.003 | 0.005 | -0.629 | 0.53 | -0.014 | 0.007 |
| D_meandurs | Agereel | 0 | 0 | -0.02 | 0.984 | 0 | 0 |
| D_meandurs | EEG_catatonique | 0.007 | 0.007 | 1.025 | 0.306 | -0.007 | 0.021 |
| D_meandurs | total_ola_equivalent | 0 | 0 | -0.205 | 0.838 | 0 | 0 |
| D_meandurs | total_fluoxetine_equivalent | -0.001 | 0.005 | -0.13 | 0.896 | -0.01 | 0.008 |
| D_meandurs | total_diazepam_equivalent | 0 | 0 | -0.67 | 0.503 | 0 | 0 |
| D_meandurs | neurodevelopmental_disorder | 0.002 | 0.009 | 0.192 | 0.848 | -0.016 | 0.019 |
| D_meandurs | neurologic_disorder | -0.003 | 0.011 | -0.281 | 0.779 | -0.025 | 0.019 |

**Table** Results of linear regression model in the psychotic subgroup between EEG features and catatonia status with covariates of age, sex, treatment (total equivalent of diazepam, olanzapine and fluoxetine), associated diagnosis (neurodevelopmental or neurological disorder). Abbreviations: Bold font corresponds to significant effects. Coef: coefficient; SD: standard deviation; CI[2.5%] CI[97.5%] to the confidence intervals at 2.5 and 97.5% and pval for p.value.
