## Supplementary Material S5 for "Neurophysiological resting-state EEG markers of catatonia in schizophrenia and mood disorders"

| dependent | names | coef | se | T | pval | CI[2.5%] | CI[97.5%] |
|---|---|---|---|---|---|---|---|
| delta | Intercept | 0.045 | 0.002 | 22.843 | 0 | 0.041 | 0.049 |
| delta | Sexe | -0.003 | 0.001 | -1.966 | 0.05 | -0.006 | 0 |
| delta | Agereel | 0 | 0 | -3.033 | 0.003 | 0 | 0 |
| **delta** | **EEG_catatonique** | **0.005** | **0.002** | **2.374** | **0.03** | **0.001** | **0.008** |
| delta | total_diazepam_equivalent | 0 | 0 | -0.079 | 0.937 | 0 | 0 |
| delta | total_ola_equivalent | 0 | 0 | 1.434 | 0.152 | 0 | 0 |
| delta | total_fluoxetine_equivalent | -0.001 | 0.001 | -1.36 | 0.174 | -0.004 | 0.001 |
| delta | neurodevelopmental_disorder | 0.002 | 0.003 | 0.868 | 0.386 | -0.003 | 0.007 |
| delta | neurologic_disorder | 0.006 | 0.003 | 2.365 | 0.018 | 0.001 | 0.011 |
| theta | Intercept | 0.024 | 0.001 | 17.441 | 0 | 0.022 | 0.027 |
| theta | Sexe | -0.002 | 0.001 | -1.586 | 0.113 | -0.004 | 0 |
| theta | Agereel | 0 | 0 | 1.884 | 0.06 | 0 | 0 |
| theta | EEG_catatonique | 0 | 0.001 | 0.019 | 0.985 | -0.003 | 0.003 |
| theta | total_diazepam_equivalent | 0 | 0 | -0.721 | 0.471 | 0 | 0 |
| theta | total_ola_equivalent | 0 | 0 | 0.85 | 0.396 | 0 | 0 |
| theta | total_fluoxetine_equivalent | 0 | 0.001 | -0.051 | 0.959 | -0.002 | 0.001 |
| theta | neurodevelopmental_disorder | -0.003 | 0.002 | -1.441 | 0.15 | -0.006 | 0.001 |
| theta | neurologic_disorder | 0.006 | 0.002 | 3.005 | 0.003 | 0.002 | 0.009 |
| alpha | Intercept | 0.036 | 0.001 | 24.926 | 0 | 0.033 | 0.039 |
| alpha | Sexe | 0 | 0.001 | 0.093 | 0.926 | -0.002 | 0.002 |
| alpha | Agereel | 0 | 0 | -2.791 | 0.005 | 0 | 0 |
| alpha | EEG_catatonique | -0.005 | 0.001 | -3.553 | 0.002 | -0.008 | -0.002 |
| alpha | total_diazepam_equivalent | 0 | 0 | 1.226 | 0.221 | 0 | 0 |
| alpha | total_ola_equivalent | 0 | 0 | -3.737 | 0 | 0 | 0 |
| alpha | total_fluoxetine_equivalent | 0 | 0.001 | 0.182 | 0.855 | -0.001 | 0.002 |
| alpha | neurodevelopmental_disorder | 0.001 | 0.002 | 0.374 | 0.708 | -0.003 | 0.004 |
| alpha | neurologic_disorder | -0.003 | 0.002 | -1.413 | 0.158 | -0.007 | 0.001 |
| beta | Intercept | 0.006 | 0 | 15.366 | 0 | 0.005 | 0.007 |
| beta | Sexe | 0.001 | 0 | 1.855 | 0.064 | 0 | 0.001 |
| beta | Agereel | 0 | 0 | 2.576 | 0.01 | 0 | 0 |
| beta | EEG_catatonique | 0 | 0 | 0.021 | 0.983 | -0.001 | 0.001 |

| | | | | | | | |
|---|---|---|---|---|---|---|---|
| beta | total_diazepam_equivalent | 0 | 0 | -0.093 | 0.926 | 0 | 0 |
| beta | total_ola_equivalent | 0 | 0 | 1.037 | 0.3 | 0 | 0 |
| beta | total_fluoxetine_equivalent | 0 | 0 | 0.521 | 0.602 | 0 | 0.001 |
| beta | neurodevelopmental_disorder | 0 | 0 | 0.039 | 0.969 | -0.001 | 0.001 |
| beta | neurologic_disorder | -0.002 | 0.001 | -2.966 | 0.003 | -0.003 | -0.001 |
| gamma | Intercept | 0.001 | 0 | 8.176 | 0 | 0.001 | 0.001 |
| gamma | Sexe | 0 | 0 | 3.489 | 0.001 | 0 | 0.001 |
| gamma | Agereel | 0 | 0 | 2.712 | 0.007 | 0 | 0 |
| gamma | EEG_catatonique | 0 | 0 | 3.146 | 0.008 | 0 | 0.001 |
| gamma | total_diazepam_equivalent | 0 | 0 | -0.98 | 0.327 | 0 | 0 |
| gamma | total_ola_equivalent | 0 | 0 | 0.482 | 0.63 | 0 | 0 |
| gamma | total_fluoxetine_equivalent | 0 | 0 | 1.844 | 0.066 | 0 | 0 |
| gamma | neurodevelopmental_disorder | 0 | 0 | 0.236 | 0.813 | 0 | 0 |
| gamma | neurologic_disorder | 0 | 0 | -0.836 | 0.404 | 0 | 0 |
| alpha_peak | Intercept | 9.834 | 0.098 | 100.041 | 0 | 9.64 | 10.027 |
| alpha_peak | Sexe | 0.132 | 0.072 | 1.829 | 0.068 | -0.01 | 0.273 |
| alpha_peak | Agereel | -0.01 | 0.002 | -4.88 | 0 | -0.014 | -0.006 |
| alpha_peak | EEG_catatonique | -0.248 | 0.095 | -2.603 | 0.009 | -0.435 | -0.061 |
| alpha_peak | total_diazepam_equivalent | -0.001 | 0.001 | -0.581 | 0.561 | -0.003 | 0.002 |
| alpha_peak | total_ola_equivalent | 0 | 0.001 | -0.001 | 0.999 | -0.001 | 0.001 |
| alpha_peak | total_fluoxetine_equivalent | 0.056 | 0.053 | 1.05 | 0.294 | -0.048 | 0.16 |
| alpha_peak | neurodevelopmental_disorder | 0.315 | 0.125 | 2.519 | 0.012 | 0.069 | 0.56 |
| alpha_peak | neurologic_disorder | -0.308 | 0.132 | -2.334 | 0.02 | -0.568 | -0.049 |
| C_occurrences | Intercept | 3.092 | 0.218 | 14.161 | 0 | 2.664 | 3.521 |
| C_occurrences | Sexe | -0.179 | 0.158 | -1.132 | 0.258 | -0.49 | 0.132 |
| C_occurrences | Agereel | -0.002 | 0.004 | -0.513 | 0.608 | -0.011 | 0.006 |
| C_occurrences | EEG_catatonique | 0.046 | 0.204 | 0.226 | 0.821 | -0.354 | 0.446 |

| | | | | | | | |
|---|---|---|---|---|---|---|---|
| C_occurrences | total_ola_equivalent | -0.002 | 0.002 | -1.102 | 0.271 | -0.005 | 0.001 |
| C_occurrences | total_fluoxetine_equivalent | 0.07 | 0.124 | 0.565 | 0.573 | -0.173 | 0.313 |
| C_occurrences | total_diazepam_equivalent | 0.002 | 0.002 | 0.9 | 0.369 | -0.003 | 0.007 |
| C_occurrences | neurodevelopmental_disorder | 0.257 | 0.275 | 0.934 | 0.351 | -0.284 | 0.798 |
| C_occurrences | neurologic_disorder | 0.002 | 0.292 | 0.008 | 0.994 | -0.572 | 0.576 |
| C_timecov | Intercept | 0.144 | 0.014 | 10.696 | 0 | 0.118 | 0.171 |
| C_timecov | Sexe | -0.006 | 0.01 | -0.604 | 0.546 | -0.025 | 0.013 |
| C_timecov | Agereel | 0 | 0 | -0.673 | 0.501 | -0.001 | 0 |
| C_timecov | EEG_catatonique | 0.012 | 0.013 | 0.967 | 0.334 | -0.013 | 0.037 |
| C_timecov | total_ola_equivalent | 0 | 0 | -0.726 | 0.468 | 0 | 0 |
| C_timecov | total_fluoxetine_equivalent | 0.005 | 0.008 | 0.716 | 0.474 | -0.01 | 0.021 |
| C_timecov | total_diazepam_equivalent | 0 | 0 | 0.85 | 0.396 | 0 | 0 |
| C_timecov | neurodevelopmental_disorder | 0.012 | 0.017 | 0.731 | 0.465 | -0.021 | 0.046 |
| C_timecov | neurologic_disorder | -0.003 | 0.018 | -0.18 | 0.857 | -0.039 | 0.032 |
| C_meandurs | Intercept | 0.042 | 0.001 | 33.642 | 0 | 0.04 | 0.045 |
| C_meandurs | Sexe | 0 | 0.001 | -0.085 | 0.932 | -0.002 | 0.002 |
| C_meandurs | Agereel | 0 | 0 | -0.756 | 0.45 | 0 | 0 |
| C_meandurs | EEG_catatonique | 0.003 | 0.001 | 2.176 | 0.03 | 0 | 0.005 |
| C_meandurs | total_ola_equivalent | 0 | 0 | -0.703 | 0.482 | 0 | 0 |
| C_meandurs | total_fluoxetine_equivalent | 0.001 | 0.001 | 0.701 | 0.483 | -0.001 | 0.002 |
| C_meandurs | total_diazepam_equivalent | 0 | 0 | 0.438 | 0.662 | 0 | 0 |
| C_meandurs | neurodevelopmental_disorder | 0 | 0.002 | 0.163 | 0.87 | -0.003 | 0.003 |
| C_meandurs | neurologic_disorder | 0 | 0.002 | -0.079 | 0.937 | -0.003 | 0.003 |
| D_occurrences | Intercept | 4.245 | 0.213 | 19.945 | 0 | 3.827 | 4.663 |
| D_occurrences | Sexe | 0.212 | 0.154 | 1.371 | 0.171 | -0.092 | 0.515 |
| D_occurrences | Agereel | -0.007 | 0.004 | -1.591 | 0.112 | -0.015 | 0.002 |

| | | | | | | | |
|---|---|---|---|---|---|---|---|
| D_occurrences | EEG_catatonique | -0.076 | 0.199 | -0.383 | 0.702 | -0.466 | 0.314 |
| D_occurrences | total_ola_equivalent | -0.001 | 0.001 | -0.508 | 0.611 | -0.004 | 0.002 |
| D_occurrences | total_fluoxetine_equivalent | -0.211 | 0.121 | -1.751 | 0.081 | -0.449 | 0.026 |
| D_occurrences | total_diazepam_equivalent | -0.002 | 0.002 | -0.706 | 0.48 | -0.006 | 0.003 |
| D_occurrences | neurodevelopmental_disorder | -0.039 | 0.268 | -0.144 | 0.885 | -0.566 | 0.488 |
| D_occurrences | neurologic_disorder | -0.188 | 0.285 | -0.661 | 0.509 | -0.748 | 0.371 |
| D_timecov | Intercept | 0.265 | 0.025 | 10.446 | 0 | 0.215 | 0.315 |
| D_timecov | Sexe | 0.016 | 0.018 | 0.871 | 0.384 | -0.02 | 0.052 |
| D_timecov | Agereel | 0 | 0.001 | -0.559 | 0.577 | -0.001 | 0.001 |
| D_timecov | EEG_catatonique | -0.016 | 0.024 | -0.677 | 0.498 | -0.063 | 0.03 |
| D_timecov | total_ola_equivalent | 0 | 0 | 0.015 | 0.988 | 0 | 0 |
| D_timecov | total_fluoxetine_equivalent | 0.006 | 0.014 | 0.384 | 0.701 | -0.023 | 0.034 |
| D_timecov | total_diazepam_equivalent | 0 | 0 | -1.221 | 0.223 | -0.001 | 0 |
| D_timecov | neurodevelopmental_disorder | -0.028 | 0.032 | -0.87 | 0.385 | -0.091 | 0.035 |
| D_timecov | neurologic_disorder | 0.004 | 0.034 | 0.124 | 0.901 | -0.062 | 0.071 |
| D_meandurs | Intercept | 0.059 | 0.005 | 10.68 | 0 | 0.048 | 0.07 |
| D_meandurs | Sexe | 0 | 0.004 | 0.049 | 0.961 | -0.008 | 0.008 |
| D_meandurs | Agereel | 0 | 0 | 0.159 | 0.873 | 0 | 0 |
| D_meandurs | EEG_catatonique | -0.001 | 0.005 | -0.274 | 0.784 | -0.011 | 0.009 |
| D_meandurs | total_ola_equivalent | 0 | 0 | 0.247 | 0.805 | 0 | 0 |
| D_meandurs | total_fluoxetine_equivalent | 0.002 | 0.003 | 0.611 | 0.541 | -0.004 | 0.008 |
| D_meandurs | total_diazepam_equivalent | 0 | 0 | -0.996 | 0.32 | 0 | 0 |
| D_meandurs | neurodevelopmental_disorder | -0.002 | 0.007 | -0.224 | 0.823 | -0.015 | 0.012 |
| D_meandurs | neurologic_disorder | 0.005 | 0.007 | 0.741 | 0.459 | -0.009 | 0.02 |
