## Supplementary Material S4 for "Neurophysiological resting-state EEG markers of catatonia in schizophrenia and mood disorders"

| | Without diazepam | | Tests | |
|---|---|---|---|---|
| N=349 | Catatonia (N=51) | Without Catatonia (N=298) | Z or X² | p |
| Age at EEG, M ± SD | 52±19 | 42.5±19 | 3.430 | **0.001** |
| Sex (F/M) | 34 (66) /17 (33) | 137 (46) /161 (54) | 6.65 | **0.009** |
| Psychotic disorder N(%) | 32 (63) /19 (37) | 201 (67) / 97 (33) | 0.25 | 0.61 |
| Mood disorder N(%)<br>Bipolar disorder N(%) | 28 (55) / 23 (45)<br>12 (23) / 39 (76) | 156 (52) / 142 (48)<br>105 (35) / 93 (31) | 0.034<br>2.1 | 0.85<br>0.14 |
| Neurodevelopmental disorder N(%) | 6 (12) / 45 (88) | 24 (8) / 274 (92) | 0.36 | 0.54 |
| Neurologic disorder N(%) | 11 (21) / 40 (78) | 20 (7) / 278 (93) | 10.1 | **0.0014** |
| Olanzapine equivalent dosage (mg), M± SD | 22.2 ± 29.8 | 24 ± 42.1 | -0.375 | 0.708 |
| Fluoxetine equivalent dosage (40 mg), M± SD | 0.42± 0.66 | 0.31± 0.65 | 1.26 | 0.260 |

**Table 1**. Description of the study population without diazepam. Bold font indicates statistically significant effects. Comparisons were made using either the Z-score proportion test or the Chi$^2$, as appropriate. Results are presented as mean ± standard deviation (M ± SD) or as absolute numbers (N) with corresponding percentages
