## Supplementary Material S3 for "Neurophysiological resting-state EEG markers of catatonia in schizophrenia and mood disorders"

| | total_ola_equivalent | total_fluoxetine_equivalent | total_diazepam_equivalent |
|---|---|---|---|
| total_diazepam_equivalent | 0.6745 | 0.6758 | 0 |
| total_fluoxetine_equivalent | 0.0513 | 0 | 0.6758 |
| total_ola_equivalent | 0 | 0.0513 | 0.6745 |

Table. P.value of the correlation matrix between treatment equivalent dosage
